# Cycle-based neurostimulation for control of circadian brain network dynamics in human epilepsy

**DOI:** 10.64898/2026.11.19.26368528

**Authors:** Krishnakant V. Saboo, Joline M. Fan, Ehsan Tadayon, Edward F. Chang, Vikram R. Rao, Ankit N. Khambhati

## Abstract

Biological rhythms modulate symptoms of brain disorders, yet brain stimulation therapies largely overlook the temporal organization of pathological brain states. Harnessing these rhythms could improve therapeutic efficacy. Here, we show that circadian coordination of hippocampal network connectivity is a biomarker of therapeutic response to responsive neurostimulation in epilepsy. Using longitudinal electrocorticography from individuals with bitemporal epilepsy, we find that alignment of circadian connectivity rhythms between the left and right hippocampus predicts seizure reduction. The daily distribution of stimulation also predicts how these rhythms evolve over months. We capture these dynamics using a nonlinear Kuramoto oscillator model and demonstrate that optimizing stimulation timing *in silico* can improve predicted circadian network alignment. Our findings establish a cycle-based, control-theoretic framework for modelling and potentially reshaping brain network rhythms, offering new opportunities for personalized neurostimulation in epilepsy and other neurological and psychiatric disorders.

## Introduction

Epilepsy operates on a clock. Epileptic seizures, like many neurological and psychiatric symptoms such as mood, anxiety, and pain, are not entirely random events; they are modulated by rhythmic brain activity spanning circadian and multidien (multi-day) timescales^1,2^. Yet, existing closed-loop brain stimulation systems treat pathological activity as though they were temporally isolated events: detect an abnormal pattern, deliver stimulation, and repeat. Although effective for some patients, this reactive paradigm does not explicitly account for cyclical brain activity that influence emission rates of pathologic events or state-dependent effects of stimulation^3–6^. Circadian rhythms are believed to create recurring changes in brain excitability that influence both seizure likelihood and therapeutic effects of stimulation^7,8^. Cycle-aware brain stimulation offers the possibility of moving beyond interrupting discrete pathological events towards reshaping the underlying rhythms that modulate their occurrence^9^.

In drug-resistant focal epilepsy, a striking temporal disconnect separates the delivery of closed-loop stimulation from the emergence of clinical benefit. The NeuroPace Responsive Neurostimulation (RNS) System delivers brief bursts of electrical stimulation in response to epileptic activity detected through intracranial electrocorticography (ECoG). While RNS benefits many patients, treatment responses remain variable, seizure reduction can take years to emerge, and complete seizure freedom is uncommon^10^. Originally thought to terminate incipient seizures, RNS is increasingly thought to exert its therapeutic effects through repeated bursts of stimulation that cumulatively alter brain network dynamics^11^. Circadian rhythms offer a potential link between these timescales, as stimulation bursts are distributed across daily cycles of fluctuating brain states. Yet, whether the circadian patterning of stimulation bursts reinforces beneficial network plasticity and ultimately drives clinical improvement remains poorly understood^12,13^. Bridging this knowledge gap requires computational models that link stimulation timing, longitudinal network dynamics, and treatment response, laying the foundation for cycle-guided control of brain activity.

Development of such models presents three challenges. First, how circadian cycles shape seizure likelihood within the epileptic brain network remains poorly understood. Functional connectivity (FC) of the bilateral hippocampi, which captures synchronized brain activity that reflects communication spanning these structures, has been established as a candidate biomarker of seizure likelihood^14^ and long-term seizure reduction^12^. It remains unclear whether FC itself follows circadian rhythms, how these rhythms align across brain regions, and whether their coordination reflects treatment response. Second, the effects of time-dependent stimulation on FC reorganization and treatment response must be resolved. Previous studies examined how stimulation effects depend on seizure risk states^3,15^ but have not established how the daily patterning of stimulation bursts shapes circadian variation of FC. Third, capturing the nonlinear evolution of network dynamics demands interpretable computational models capable of learning from sparse longitudinal recordings available in clinical practice.

Here, we develop a cycle-guided computational framework integrating circadian brain network dynamics, daily stimulation patterns, and clinical outcomes in people with bilateral mesial temporal lobe (bitemporal) epilepsy. We first examine how circadian cycling of FC provides a dynamic marker of treatment response. We next investigate whether stimulation timing contributes to the long-term evolution of these circadian cycles. Lastly, we develop a dynamical oscillator model to explore how stimulation could be tuned to reshape circadian network dynamics and design individualized treatment strategies *in silico*. Together, we demonstrate a novel approach for moving beyond reactive, event-triggered stimulation towards deliberate, directed modulation of circadian brain network dynamics in epilepsy.

## Results

We studied 38 adults (21 males; Supplementary Table S1) with bilateral mesial temporal lobe (bitemporal) epilepsy who were implanted chronically with the RNS System with bilateral hippocampal depth leads that recorded neural activity (Fig. 1a). During routine clinical encounters, monthly patient-reported seizure frequency was compared to seizure frequency during a 3-month pre-implantation baseline period. Analysis was restricted to visits for which records were available.

**Figure 1:**
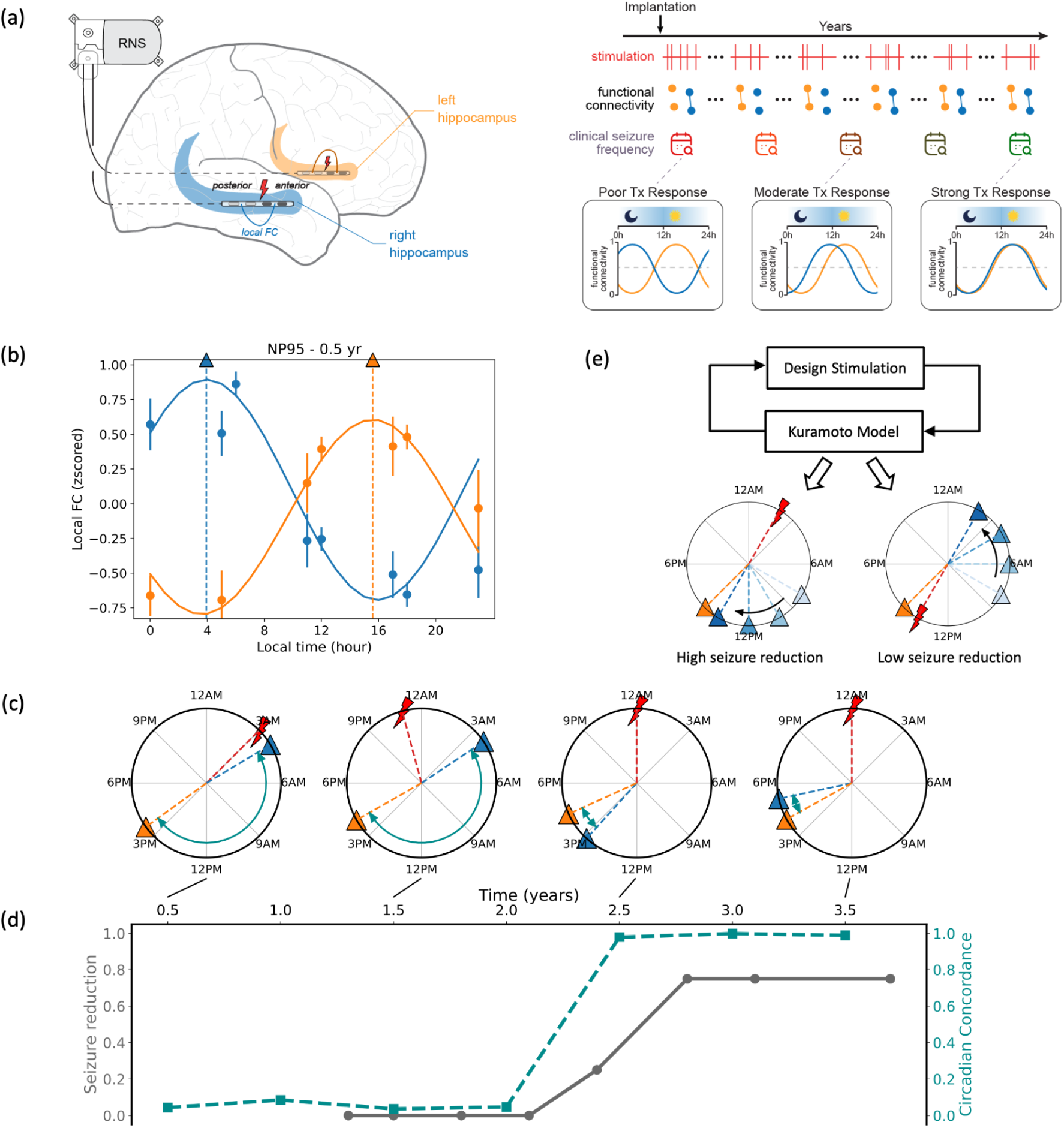
Study overview. (a) Functional connectivity (FC) and stimulation data collected over days from bilateral hippocampal RNS implants were time-matched to clinical seizure frequency for biomarker discovery. (b) FC circadian cycle estimation from FC measures aggregated over 6-month windows. Example data for one six-month window (epoch) of a participant. (c) Peak hour of circadian cycle in alpha band for left and right hippocampus and stimulation for the participant NP95 over different 6-month epochs. (d) Circadian concordance, a measure of alignment between bilateral hippocampal FC circadian cycles, and seizure reduction for the participant NP95 over the entire duration of their recording. (e) Kuramoto model of oscillators describes the modulation of circadian peak and circadian concordance from stimulation.

### Network concordance of circadian cycles

To measure time-dependent, intrahippocampal communication, we computed a functional connectivity (FC) metric called the phase locking value (PLV) between RNS contacts in the anterior and the posterior hippocampus. Left- and right-sided hippocampal FC was obtained for 99,349 scheduled electrocorticography (ECoG) recordings collected by the RNS device over 66,732 participant-days. Recordings were acquired at participant-specific predetermined intervals (Fig. S1).

We modelled circadian dynamics in left and right hippocampal FC separately using Lomb-Scargle (LS) spectral analysis, a method that detects repeating patterns in irregularly sampled timeseries (Fig. 1b). LS models with a 24-hour period were fitted to FC values over time to obtain the amplitude and phase of network circadian dynamics. LS models were fitted independently for each frequency band and channel pair within non-overlapping six-month windows (epochs) anchored to the date of implantation. Across the cohort, network circadian cycles were evident across frequency bands, with peak phases spanning the 24-hour clock (Fig S2).

We anticipated that the circadian cycling of FC in the left and right hippocampi would rise and fall at the same times of the day. Surprisingly, in an example individual, we discovered that the left- and right-sided circadian cycles were initially distinct and dynamically reorganized during RNS treatment (Fig. 1c). Specifically, circadian cycling within the left and right hippocampi became more aligned, or *concordant*, with greater seizure reduction (Fig. 1d). We hypothesized that (1) seizure reduction is linked to circadian reorganization of hippocampal FC, and (2) circadian reorganization is driven in part by the relative timing of stimulation bursts over the course of the day (Fig. 1e). To test these hypotheses, we evaluated circadian cycle concordance as a candidate biomarker of treatment response across the study cohort.

### Biomarker of ongoing seizure reduction

Longitudinal seizure outcomes varied across participants (Fig. 2a). Progressive seizure reduction was observed across the cohort over multiple years, but seizure frequency fluctuated over time. For example, participants demonstrated periods of worsening seizure frequency following intervals of near-complete seizure freedom. Given these fluctuations, we posit that there may be biological signals that underlie the RNS treatment course. We therefore asked whether hippocampal network circadian dynamics could predict ongoing seizure reduction. We found that greater alignment of circadian cycles between the left and right hippocampus differentiated visits with high seizure reduction (> 0.90) from visits with low seizure reduction (< 0.50) (Mann-Whitney U test *P* = 0.0009; Fig. 2b).

**Figure 2:**
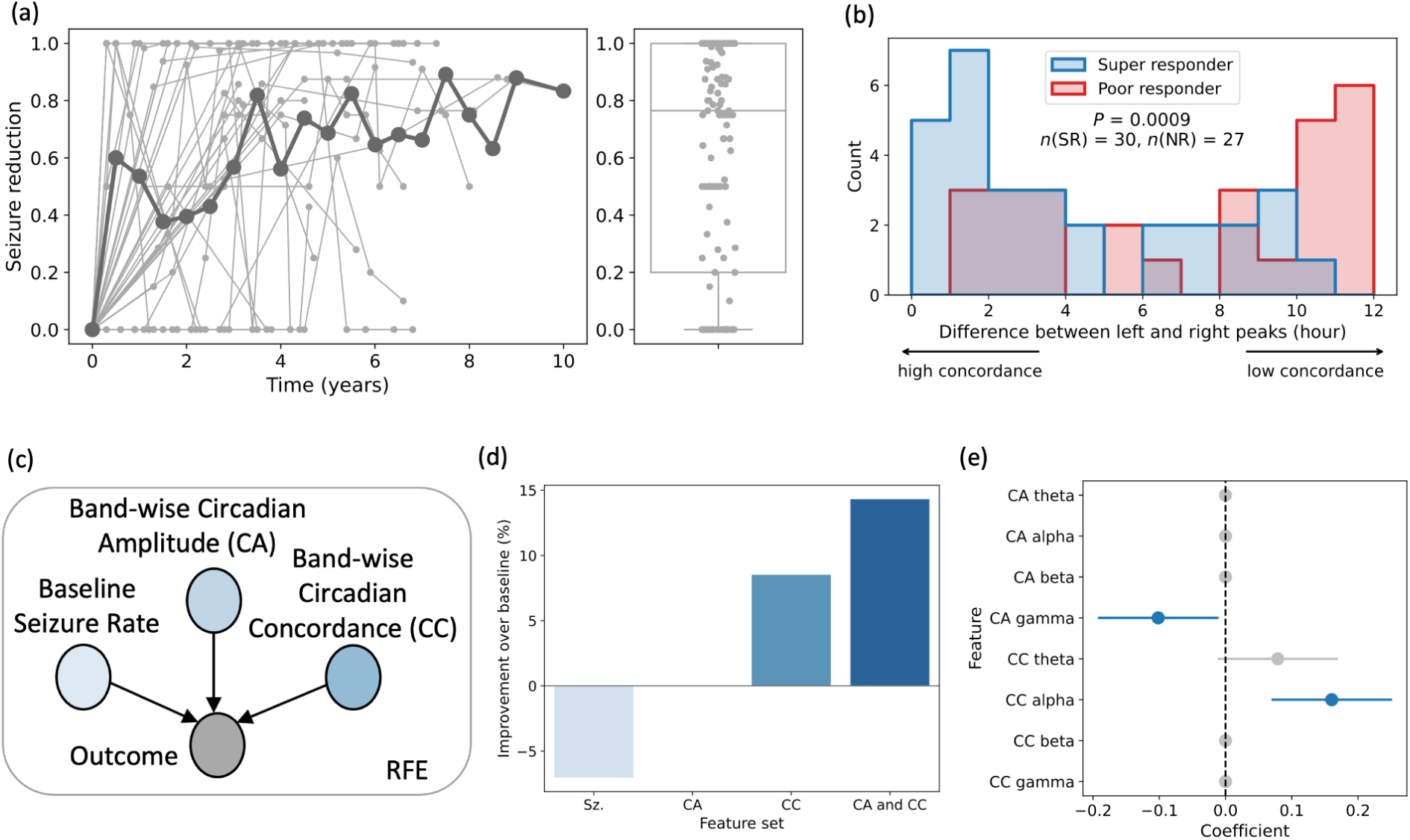
Circadian concordance in alpha is a biomarker of ongoing seizure reduction. (a) Outcome, i.e., seizure reduction, over time for all the participants (bold line shows the mean). (b) Comparison by treatment outcome of difference between left and right network circadian cycle peaks in the alpha band. Hourly difference values were compared using Mann-Whitney U test. (c) Outcome prediction using band-specific circadian amplitude (CA) and circadian concordance (CC), alongside log baseline seizure rate as a covariate, with important features identified through recursive feature elimination (RFE). Leave-one-participant-out (LOPO) cross-validation was performed. (d) Circadian features improve outcome prediction over population mean baseline. (e) Beta weights of prediction model across LOPO CV folds show that circadian concordance in alpha is a significant predictor of outcome. Features that were eliminated during RFE have a coefficient of 0 (shown in shaded gray).

To further investigate circadian network dynamics as a predictor of treatment response, we constructed multivariate regression models to predict longitudinal seizure reduction from time-matched circadian network features. Model performance was evaluated using leave-one-participant-out cross-validation (LOPO CV), and mean prediction errors were computed on held-out test sets (Methods) (Fig. 2c).

Circadian features improved the prediction of time-matched single participant treatment response by 14.3% above chance (Fig. 2d). We observed weakened gamma-band circadian amplitude and strengthened alpha-band circadian concordance as predictors of longitudinal seizure reduction (Fig. 2e). Higher circadian concordance in the alpha band was associated with improved ongoing seizure reduction (Fig S3). We rigorously assessed the robustness of this finding across stricter detection criteria of circadian cycles (Fig. S4). Further, the connectivity-based marker of circadian concordance outperformed power-based markers of circadian concordance in predicting clinical seizure reduction by 57.5% (Fig. S5). Henceforth, circadian concordance (CC) in the remainder of this paper refers specifically to concordance in the alpha frequency band due to its strong predictive effect.

### Clustering of peak hours of network circadian cycles

We next asked whether the circadian rhythms spanning the hippocampal network have generalizable and stable organization across participants and time. Pooling circadian peak hours across participants, left and right hemispheres, and epochs, we observed a markedly non-uniform distribution (Kuiper test, *n* = 466, *P* = 5.2e-6) (Fig. 3a), with peak hours forming one distinct nocturnal cluster (centered at 3:39AM ± 4.1 mins) and another distinct diurnal cluster (centered at 3:02PM ± 3.5 mins).

**Figure 3:**
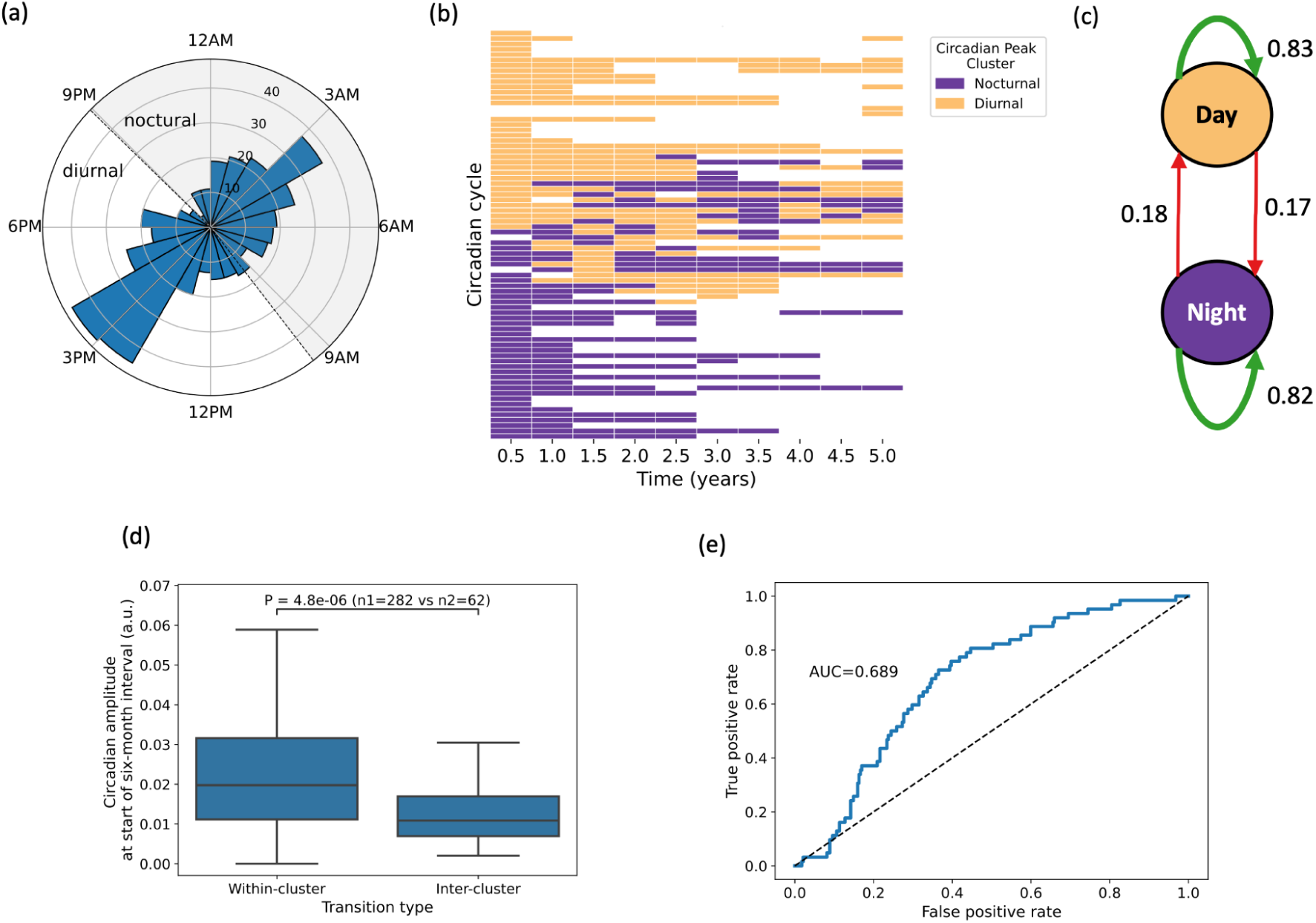
Peak hour of circadian cycle in alpha forms clusters and changes between and within the cluster over time. (a) Distribution of peak hour of circadian cycle alpha across all participants and six-month epochs. Peaks clustered around diurnal and nocturnal attractors. Boundaries between the clusters are shown in dotted lines. (b) Cluster membership of circadian cycle peak hour over 5 years for different participants and hemispheres. Transition heatmap for all 10 years of available data provided in Supplement. (c) Markov model with transition probability within and between diurnal and nocturnal clusters. (d) Circadian amplitude at the start of epoch is significantly different between within- and inter-cluster transitions. (e) Receiver operating characteristic (ROC) curve for a logistic regression model that predicts transition type based on circadian amplitude at the start of the epoch. LOPO cross-validation was performed.

We investigated the longitudinal stability of cluster membership within each participant-hippocampus in six-month epochs and found that cycles tended to remain within the same nocturnal or diurnal cluster to which it was originally assigned at the beginning of RNS treatment (Fig. 3b). Transitions between clusters were relatively infrequent and could occur in either direction (diurnal-to-nocturnal or nocturnal-to-diurnal), with individual cycles potentially visiting both clusters multiple times over the recording period (Fig. S6). State transition analysis revealed that cycles were approximately four times more likely to remain within their current cluster than to transition to the other (Fig. 3c; within-cluster transition probability: 0.83 and 0.82 for diurnal and nocturnal states, respectively; inter-cluster transition probability: 0.17 and 0.18). The near symmetry of transition probabilities across clusters indicates that neither phase state is inherently more stable than the other.

Finally, we examined whether intrinsic properties of a circadian cycle could prospectively predict the type of transition it would undergo. Circadian amplitude was significantly lower in cycles that subsequently underwent inter-cluster transitions compared to those that remained within the same cluster (2-sample t-test; *n*₁ = 282, *n*₂ = 62; *P* = 4.8e-6; Fig. 3d). A logistic regression model trained to predict transition type from cycle amplitude and phase achieved an area under the receiver operating characteristic curve (AUROC) of 0.689 in leave-one-participant-out cross-validation (Fig. 3e), confirming the prospective utility of this feature.

Taken together, these findings reveal that hippocampal network circadian cycles organize around nocturnal or diurnal *attractors*. Low-amplitude cycles are more susceptible to transitions between clusters centered at these attractors. Crucially, cycle amplitude can identify, six months in advance, those participants whose circadian concordance is likely to undergo a substantial reorganization, opening a potential window for prospective therapeutic intervention.

### Circadian patterning of stimulation events

We next sought to test whether RNS therapy drives changes in circadian cycles underlying treatment response. Based on our finding that the circadian rhythm predicts treatment response, we reasoned that the temporal patterning of stimulation events over the course of the day may also relate to seizure control. Total stimulation count did not predict treatment response (Fig. S7). The stimulation profile curve, which quantifies the normalized distribution of hourly stimulation event counts per participant, differentiated super responders (seizure reduction > 90%) from poor responders (seizure reduction < 50%; Fig. 4a). After aligning each participant’s stimulation profile curve to their peak hour, super responders exhibited a temporally narrower stimulation profile compared to poor responders (2-sample t-test at each hour; *n_1_* = 8, *n_2_* =10, *P_uncorrected_* < 0.05 between 1h and 7h relative to stimulation peak hour).

**Figure 4:**
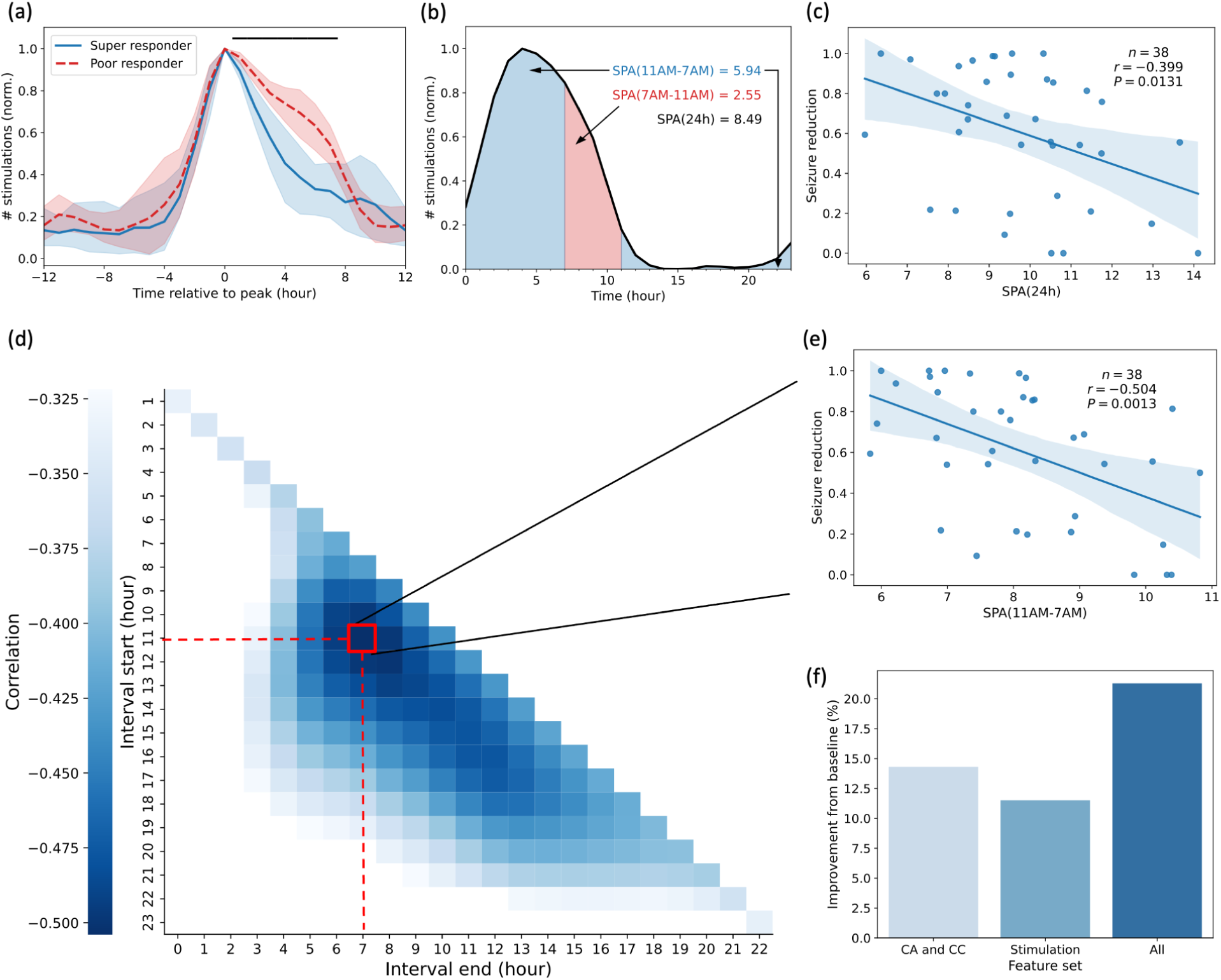
Stimulation profile curves predict seizure reduction. (a) Difference in circadian profile of normalized stimulation between super responders (seizure reduction > 0.9) and poor responders (seizure reduction < 0.5). Stimulation profiles were aligned such that maximum stimulation occurred at 0h within each participant. Solid black line denotes hours with a significant difference in stimulation between responder groups. (b) Computation of stimulation profile area (SPA) over 24 hours and for intervals for an example participant. (c) Lower SPA(24h) predicts larger seizure reduction. (d) Correlation between average seizure reduction and SPA for different intervals. Significant correlations are shown (*p* < 0.05; uncorrected due to non-independent, overlapping intervals of the data). (e) Correlation between average seizure reduction and SPA(11AM–7AM). (f) LOPO CV model performance for outcome prediction using band-specific circadian amplitude (CA) and circadian concordance (CC), and stimulation features alongside log baseline seizure rate as a covariate. Important features identified through RFE.

To quantify the temporal concentration of stimulation, we defined stimulation profile area (SPA) as the area under the normalized stimulation profile curve (Fig. 4b). Intuitively, lower SPA indicates stimulation is concentrated within fewer hours of the day, whereas higher SPA indicates that stimulation remains elevated over a broader portion of the day. Thus, SPA is sensitive to the relative temporal distribution of stimulation over a 24-hour period rather than the amount of stimulation.

SPA over 24 hours, defined as SPA(24h), was negatively correlated with average seizure reduction across participants (*n* = 38, *r* = −0.399, *P* = 0.013; Fig. 4c), suggesting that participants with more temporally concentrated stimulation profiles over a 24-hour period experienced greater seizure reduction. We further tested whether stimulation during particular times of day was more strongly associated with treatment response. SPA was computed for all hourly intervals spanning a 24-hour period and correlated with average seizure reduction (Fig. 4d). A systematic pattern emerged: greater seizure reduction was associated with stimulation concentrated into a narrower portion of the day. Specifically, participants with lower SPA within the window beginning at 11 AM and ending at 7 AM the following morning experienced greater seizure reduction (*n* = 38, *r* = −0.504, *P_uncorrected_* = 0.0013; Fig. 4e). This finding indicates that less broadly distributed stimulation across afternoon, evening, and overnight hours is associated with better treatment response.

We compared the utility of stimulation features, i.e., SPA and stimulation peak phase, with network circadian dynamics features for predicting seizure reduction. Multivariate regression models were trained with RFE and evaluated using LOPO CV (Fig. S8). Network circadian features alone outperformed stimulation features in predicting longitudinal seizure reduction (Fig. 4f). Combining stimulation and circadian network features yielded the strongest overall model, reducing prediction error by 21.3% relative to the baseline mean predictor.

Collectively, these results demonstrate that the daily timing of RNS stimulation carries important information about treatment response. Greater seizure reduction was associated with stimulation concentrated around a narrower daily peak, rather than sustained across much of the day, particularly outside the afternoon, evening, and overnight hours. Circadian organization of the stimulation profile curve captures therapeutic information complementary to that encoded in hippocampal network dynamics.

### Bidirectional control of network circadian cycles

What drives reorganization of the circadian cycle and determines the change in peak phase? An answer to this question would provide mechanistic insight into how effective long-term neurostimulation re-aligns circadian rhythms across the hippocampi. We trained a multivariate regression model to forecast directional changes in the peak hour of the circadian cycle observed six months ahead (Fig. 5a). Models incorporated the initial state of the circadian cycle and stimulation features during the ensuing six months as predictors.

**Figure 5:**
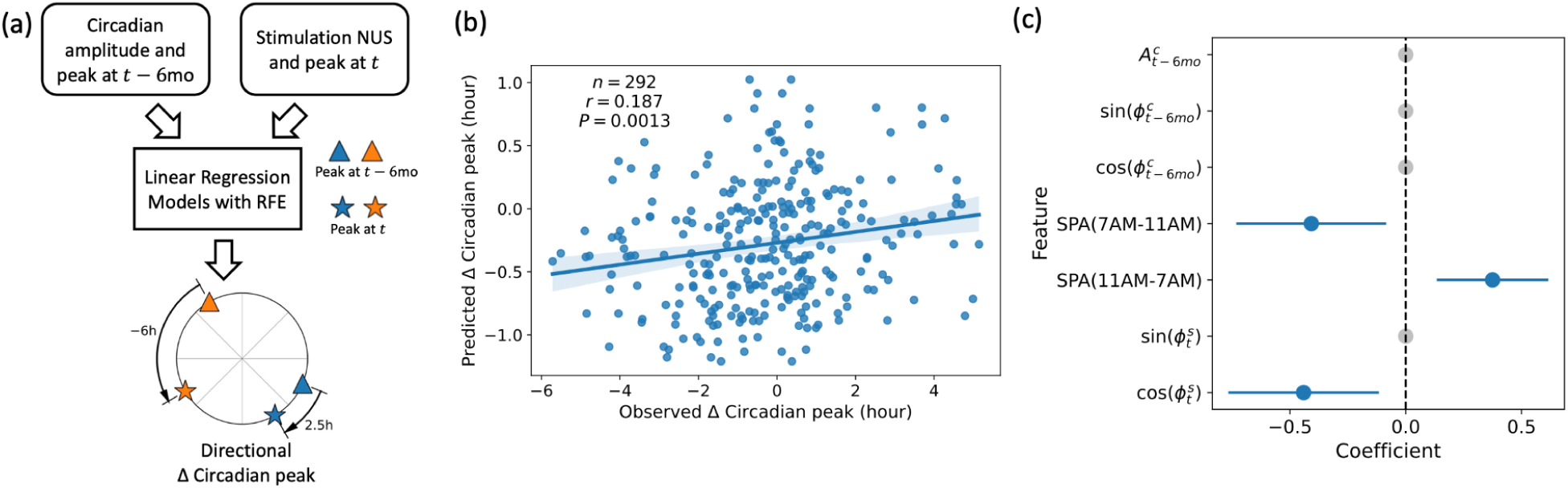
Stimulation modulates the peak hour of circadian cycles. (a) Linear models predicted the directional change in circadian cycle peak between successive 6-month epochs based on circadian cycle characteristics at the start of the epoch (t−6mo) and stimulation during the epoch (t). (b, c) LOPO CV model performance for predicting directional change in circadian peak and beta weight across folds. Features that were eliminated during RFE have a coefficient of 0 (shown in shaded gray). Abbreviation: RFE – recursive feature elimination;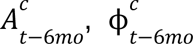 – circadian cycle amplitude and peak phase at the start of the six-month epoch; 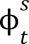 – stimulation peak phase during the six-month epoch.

Temporal patterning of stimulation forecasted the directional change in peak hour (LOPO CV; *n* = 292, *r* = 0.187, *P* = 0.0013; Fig. 5b, Fig. 5c). Stimulation that was temporally concentrated in the morning predicted an anti-clockwise shift in circadian peak hour, whereas stimulation distributed broadly across the afternoon, evening, and night predicted a clockwise shift.

This finding demonstrates that the timing of neurostimulation actively shapes hippocampal network circadian dynamics over months, supporting the hypothesis of a stimulation mechanism operating at circadian timescales. The opposing directional effects of morning versus non-morning stimulation raise the intriguing possibility that stimulation could be deliberately designed to steer circadian peak hours toward bilateral alignment and thereby toward greater seizure reduction. We explore this possibility in the following two sub-sections.

### Kuramoto oscillators model long-term dynamics of network circadian cycles

We sought to develop a mechanistic mathematical model capable of unifying the interactions between network circadian dynamics, stimulation patterning, and nocturnal/ diurnal attractors. Such a model would move beyond empirical correlations toward a mathematical description of network circadian evolution under the influence of stimulation.

We modelled the system using a Kuramoto oscillator framework, for three principal reasons. First, hippocampal network circadian cycles follow oscillatory dynamics. Second, interactions between oscillators may drive changes in the circadian peak hour, and can be parameterized by coupling constants in the Kuramoto model. Third, by expressing these interactions as a function of the phase difference between oscillators, the model can account for the clustering of peak hours around the two attractors (Fig. 3).

The model comprised five oscillators, each with a 24-hour period: (i) the left hippocampal FC circadian cycle, (ii) the right hippocampal FC circadian cycle, (iii) stimulation, (iv) a diurnal attractor fixed at a peak phase of 3 PM, and (v) a nocturnal attractor fixed at a peak phase of 3 AM (Fig. 6a). The hippocampal network cycles were included as the primary state variables whose evolution we sought to model. Stimulation was included as a driving oscillator, given its demonstrated influence on circadian peak hour (Fig. 5). Inter-oscillator influences were parameterized through coupling constants *K_att_*, *K_brain_*, *K_lstim_*, and *K_rstim_.* Only the peak phases of the two hippocampal network cycles were treated as dynamic variables; the peak hour of the stimulation oscillator was derived from data and held constant within each six-month epoch, and the peak phases of the attractor oscillators were fixed throughout.

**Figure 6:**
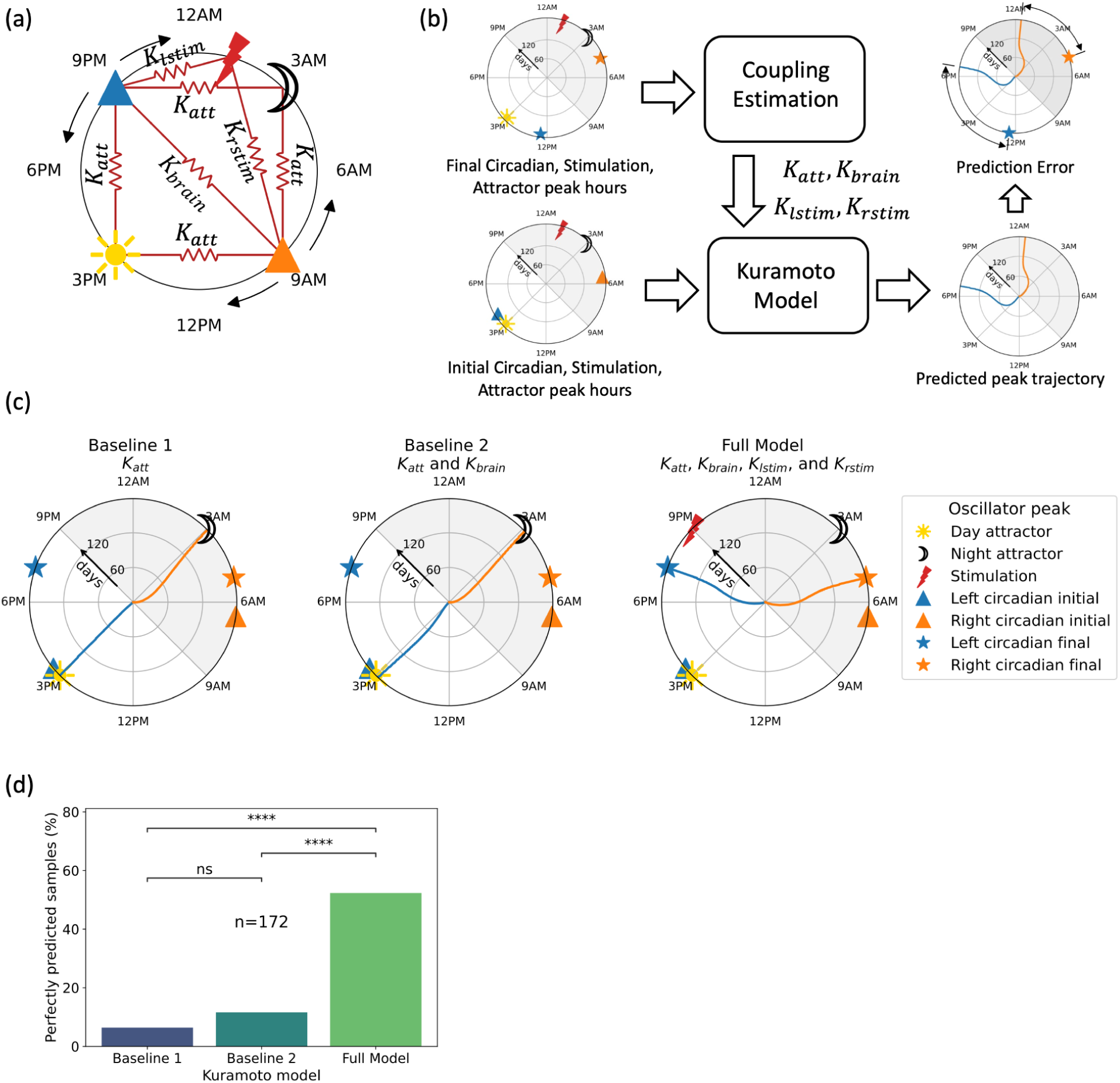
Kuramoto model captures the interactions between stimulation and circadian cycles in real data. (a) Kuramoto model with stimulation, circadian cycles, and diurnal and nocturnal attractors as oscillators. Markers represent peak phase of the oscillators and couplings are represented by springs. (b) Workflow to estimate coupling constants, predict simulated trajectory of peaks, and compute prediction error for real data. (c) Predicted trajectory of circadian peaks over six months for a real sample for baseline and full Kuramoto models with different coupling constants. Coupling values were scaled to visually emphasize the non-linearity of the trajectories. Unscaled trajectories are provided in the Supplement. (d) Percentage of samples (six-month epochs) with prediction error less than an hour for Kuramoto models with different coupling constants. Pairwise statistical comparisons from two-proportion Z-test are shown. ns: *P*≥0.05, **** *P*<0.0001.

For each six-month epoch in a participant, the Kuramoto model predicted the evolution of peak phases of circadian cycles over time based on stimulation and the circadian cycles’ peak phase at the start of the epoch. To contextualize model performance, two baseline models were also trained.

Illustrative model-predicted trajectories for a representative epoch are shown in Fig. 6c. In both baseline models, the hippocampal cycle peak hours converged toward the nearest attractor phase, irrespective of the observed outcome. In the full model (right), the predicted peak hours converged to the empirically observed terminal phases in both hemispheres, demonstrating that the inclusion of stimulation coupling was necessary to capture the actual direction and magnitude of phase change (Fig. S9).

To quantify the proportion of epochs explained with high accuracy, we computed the fraction of samples with prediction error below 1 hour. The full Kuramoto model explained 52.3% of epochs within this threshold, compared to 6.4% and 11.2% for the two baselines respectively (Two-proportion Z-test; *n* = 172; *P* < 5.9e−16) (Fig. 6d), confirming that stimulation coupling substantially improves the model’s ability to account for observed six-month phase dynamics.

Taken together, these results demonstrate that a Kuramoto oscillator model incorporating stimulation coupling can account for the majority of six-month changes in hippocampal network circadian peak hour, independent of seizure reduction (Fig. S9). This establishes the Kuramoto framework as a broadly applicable computational model of long-term network circadian evolution under responsive neurostimulation, and sets the foundation for using it as a tool for *in silico* stimulation optimization.

### In-silico optimization of stimulation

A key advantage of a mechanistic mathematical model over purely statistical approaches is its capacity for counterfactual reasoning, i.e., the ability to answer questions of the form: given a patient’s current network state, what stimulation strategy would maximize seizure reduction? We leveraged the Kuramoto framework to address this question *in silico*, focusing specifically on identifying the optimal peak hour of stimulation for a given initial configuration of hippocampal circadian cycle peak phases.

The optimization pipeline proceeded in four steps for each sample (Fig. 7a). First, a candidate peak hour of stimulation was selected. Second, multivariate regression models were used to predict the coupling constants *K_brain_*, *K_lstim_*, and *K_rstim_* for that stimulation configuration (see below). Third, the Kuramoto model was integrated forward for 180 days using the predicted coupling constants and the observed initial peak phases, and the resulting circadian concordance between left and right hemispheres was computed. Fourth, steps one through three were repeated across all 24 candidate stimulation peak hours. This procedure was applied to all available samples, and the analysis was conducted as closely as possible to a held-out test-set framework to preserve clinical applicability.

**Figure 7:**
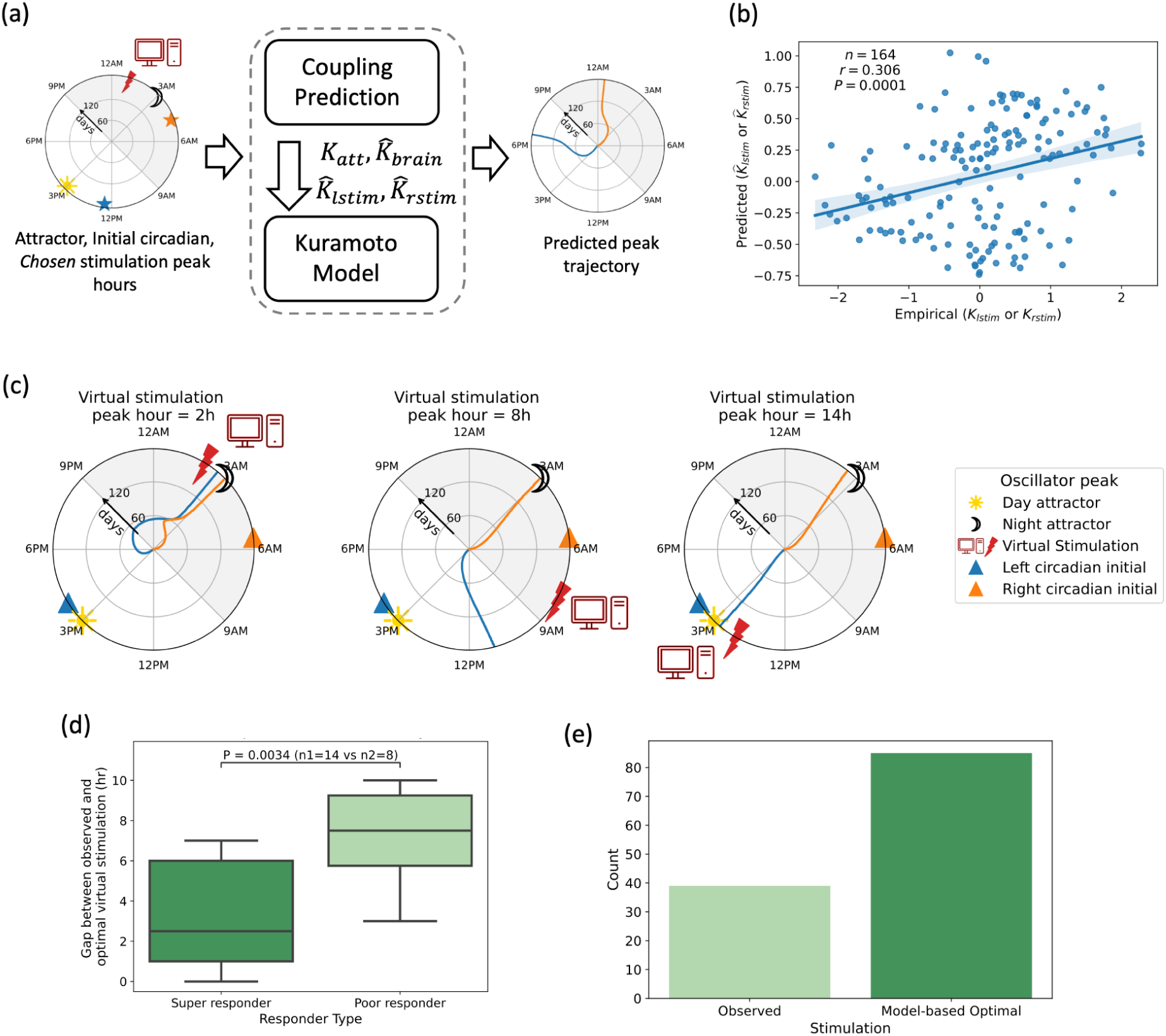
Kuramoto model-based *in silico* optimization of stimulation for improved circadian concordance. (a) Schematic for *in silico* testing of the effect of different virtual stimulations on circadian peak trajectories predicted using Kuramoto models. Virtual stimulation is shown via a computer icon and red lightning bolt. (b) LOPO cross-validated performance for predicting stimulation-cycle coupling from circadian cycle and stimulation features. (c) Effect of varying the peak hour of virtual stimulation on circadian cycles. Subplots show, for a real sample, the changes in Kuramoto model-predicted trajectory of circadian peaks over six months for different virtual stimulation peak hours. Coupling values were scaled to visually emphasize the non-linearity of the trajectories. Unscaled trajectories are provided in the Supplement. (d) Deviation between observed stimulation and optimal virtual stimulation across epochs stratified by responder status. Only epochs for which optimal virtual stimulation achieved circadian concordance > 0.95 were considered. (e) Number of epochs for which high circadian concordance (>0.95) was achieved using Kuramoto model-based optimal stimulation compared to observed stimulation.

To support the mechanistic validity of the Kuramoto framework, we predicted the coupling constants from biologically interpretable features that were not used to determine the empirical value of these constants. Multivariate models predicted Kuramoto coupling constants from initial circadian cycle and stimulation features with significantly better-than-chance accuracy under leave-one-participant-out cross-validation (*K_lstim_* and *K_rstim_* : *n* = 164, *r* = 0.306, *P* = 6.8e-5; *K_brain_* : *n* = 79, *r* = 0.505, *P* = 2e-6) (Fig. 7b; Fig. S10).

To illustrate the model’s utility for stimulation optimization, we examined predicted six-month circadian peak trajectories under three different virtual stimulation peak hours for a representative sample, in which the initial left and right hippocampal peak phases were approximately 9.5 hours apart (left: ∼3:30 PM; right: ∼6:00 AM) (Fig. 7c). When the stimulation peak was set to 2 AM, both cycles converged toward 3 AM over six months, yielding high circadian concordance. Setting the stimulation peak to 8 AM caused the left cycle to converge to approximately 11 AM and the right to 3 AM, resulting in poor concordance. A stimulation peak at 2 PM drove the left cycle toward 3 PM while the right remained near 3 AM, again producing low concordance. Among the three configurations, a 2 AM virtual stimulation peak achieved the greatest bilateral alignment and, by extension, the greatest predicted seizure reduction (Fig. S11).

To validate the *in silico* framework against empirical data, we examined whether virtually optimized stimulation aligned with observed clinical outcomes. We expected that virtually optimized stimulation would achieve high circadian concordance and closely match observed stimulation in epochs with better real-world outcomes. For each sample, the optimal virtual stimulation peak hour was defined as the candidate hour that maximized circadian concordance in that epoch. Among epochs in which virtual stimulation achieved high circadian concordance (>0.95), super-responders received stimulation significantly closer to the virtual optimum than poor responders (two-sample t-test; *n₁* = 14, *n₂* = 8; *P* = 0.0034; Fig. 7d). Furthermore, the framework reproduced the empirical association between SPA and seizure reduction without any direct exposure to this relationship during model training (Fig. S11). Together, these findings provide convergent support for the validity of the Kuramoto framework and strengthen confidence in its use for prospective stimulation design.

We therefore used the optimization framework to estimate the potential clinical benefit of Kuramoto-guided stimulation. For each sample, we compared the circadian concordance observed in real data against the maximum concordance achievable under model-optimized stimulation. Circadian concordance exceeded 0.95, indicative of near-complete bilateral alignment and associated with seizure freedom, in 85 six-month epochs under optimized stimulation, compared to 39 epochs in the observed data. This represents a greater than two-fold increase in the proportion of epochs with high concordance, suggesting that Kuramoto model-guided stimulation optimization could substantially expand the duration for which participants achieve seizure freedom relative to current clinical practice.

## Discussion

We develop a novel cycle-based control-theoretic framework for modelling the effect of brain stimulation on network circadian dynamics and leveraging this understanding for *in silico* optimization of stimulation in epilepsy. Using longitudinal ECoG recordings from individuals with bitemporal epilepsy undergoing RNS therapy, we demonstrated that the alignment of circadian network activity across the left and right hippocampus is a biomarker of ongoing seizure reduction. More temporally concentrated stimulation was associated with greater seizure reduction, revealing a previously uncharacterized relationship between the circadian patterning of stimulation and therapeutic outcome. Critically, longitudinal changes in hippocampal network circadian dynamics were predicted by the 24-hour temporal structure of stimulation. Together, these findings support a novel mechanism of RNS action operating at the circadian timescale. We formalized this mechanism using a Kuramoto oscillator model, which accounted for the six-month evolution of hippocampal network circadian dynamics under stimulation. Virtual interrogation of a population-level framework with the Kuramoto model identified optimized stimulation timing strategies that, *in silico*, doubled the proportion of epochs in which participants achieved near-complete seizure freedom relative to current clinical practice. Beyond its immediate application to hippocampal RNS in epilepsy, the proposed framework has broader relevance for the design of brain stimulation therapies across neurological and psychiatric conditions where symptoms fluctuate with regular periodicity^9,16^.

We found that bilateral alignment of circadian dynamics in the hippocampal network predicts ongoing seizure reduction. We established this biomarker by investigating the circadian dynamics of network connectivity based on the finding that FC captures long-term therapeutic response and forecasts 24-hour seizure risk^12,14^. Our study extends prior work developing biomarkers of treatment response^7,17^ by tracking circadian network dynamics and seizure reduction in parallel over years of follow-up, capturing the longitudinal heterogeneity in patient-reported outcomes that is characteristic of real-world RNS therapy^18,19^. The closest precedent for a circadian biomarker of ongoing response to brain stimulation comes from the literature on brain stimulation for psychiatric disorders. A prior study of deep brain stimulation in obsessive-compulsive disorder^16^ reported that circadian cycles of 9 Hz power tracked symptom severity and therapeutic efficacy. Our findings in epilepsy provide further evidence that the relationship between circadian neural organization and stimulation response may be a general principle applicable across neuropsychiatric conditions.

Effects of brain stimulation are thought to be state-dependent, though the precise mechanisms remain incompletely understood. Brain states related to seizure likelihood are known to modulate the effect of stimulation^3,4^ on long-term seizure control with RNS^15^. Brain states related to sleep and wakefulness, which are observable manifestations of an endogenous circadian oscillator^20^, may similarly modulate the effect of stimulation delivery^21^.

Consistent with this reasoning, we found that stimulation that was more temporally concentrated between 7AM and 11AM was associated with greater seizure reduction. This relationship has both prognostic and prescriptive value: it provides a novel indicator of likely treatment response and suggests a potential target for stimulation redesign. Relative to super-responders, poor responders tended to receive disproportionately more stimulation at night than during the day, resulting in a temporally wider distribution of stimulation across the 24-hour cycle. While RNS-delivered stimulation has not been found to disrupt sleep architecture per se^22^, excessive nocturnal stimulation may nonetheless interfere with sleep in a manner analogous to interictal epileptiform discharges, which are known to disrupt sleep^23^. Conversely, more temporally uniform stimulation across the non-morning hours may promote long-term reorganization of the epileptic network by engaging synaptic consolidation mechanisms active during sleep^24,25^, thereby contributing to the sustained therapeutic efficacy observed in RNS^12,13^. As sleep was not tracked in this retrospective cohort, we could not assess whether sleep-wake state mediates the link between stimulation timing and outcome. Future prospective studies should track sleep to disentangle its contribution from that of circadian timing.

Circadian concordance in the alpha band emerged as the strongest predictor of ongoing treatment response, consistent with prior work linking alpha band dynamics in the therapeutic effects of brain stimulation. In RNS for epilepsy, early post-implantation increases in daily alpha oscillatory power have been associated with long-term seizure reduction^7^, and in DBS for OCD circadian cycling in alpha band power predicted both symptom severity and treatment response^16^. Together, these findings suggest that alpha band activity may play a privileged role in the neural response to chronic brain stimulation, perhaps because alpha rhythms support mesoscopic communication between neuronal populations^26^. In epilepsy, bilateral hippocampal circadian misalignment may reflect partial isolation of seizure-generating tissue from broader synchronizing networks^27^, leading to attenuated circadian power in the tissue^28^ and allowing local network rhythms to drift independently. Greater concordance may therefore index a more integrated hippocampal circuit capable of distributing epileptiform activity across connected networks rather than permitting its concentration within an isolated seizure-generating zone^29^. Such alignment may also increase the reach of stimulation-evoked activity across hemispheres, enhancing the capacity of RNS to reshape pathological circuits^30,31^. An alternative interpretation is that concordance is not causal, but instead reflects successful stimulation-driven reconnection of an isolated epileptic network. Distinguishing whether circadian concordance actively mediates seizure reduction or serves as a downstream marker of network repair will be an important direction for future work.

Our principal methodological contribution is demonstrating that the Kuramoto oscillator framework can mathematically describe the longitudinal evolution of hippocampal network circadian dynamics under the influence of responsive neurostimulation. The model serves three complementary purposes: to move beyond empirical correlations toward a process-level account of how circadian network organization evolves over time; to provide mechanistic insight into how stimulation timing reshapes network oscillatory dynamics; and to furnish a predictive framework for optimizing stimulation delivery by deliberately steering circadian peak phases toward bilateral alignment.

Our study offers a platform for *in silico* optimization of the temporal patterning of neurostimulation. RNS parameter tuning is a high-dimensional problem, encompassing over 12 million possible parameter combinations with no principled basis for selection^32^ and currently relies on time-consuming empirical adjustment^33,34^. Critically, agreement between virtually optimized stimulation and verum stimulation predicted better outcomes, supporting the biological validity of the framework. By virtually identifying stimulation timing strategies that promote circadian concordance, the Kuramoto framework offers a principled approach to expediting this search and improving individual treatment outcomes. More broadly, the simplicity, neurophysiological interpretability, and established applicability of Kuramoto models across neuroscience^35^ positions this framework as a promising tool for studying the effects of brain stimulation across a wide range of timescales: from seconds to circadian and multiday periods.

Prior computational approaches have largely focused on either differential equation-based models or probabilistic state-space models^36,37^. Differential equation-based approaches, including Epileptor^38^, Virtual Brain^39,40^, and related biophysical models^41^ have described fast seizure dynamics. Probabilistic state-space models provide a complementary view by capturing neural variability and autoregressive dynamics, including the acute neural effects of stimulation^42^ and multiday cycling for seizure forecasting^43^. These models characterize dynamics at either the seconds or multiday timescale and are primarily concerned with the evolution of seizure risk rather than the network reorganization that underlies chronic treatment effectiveness. Our approach is distinct. We employ non-linear coupled differential equations to model how stimulation delivery interacts with circadian hippocampal network organization over months-long timescales. In doing so, the Kuramoto framework bridges the gap between the fast dynamics captured by seizure-onset models and the slow multiday trends captured by state-space forecasting approaches, providing a principled account of how chronic stimulation progressively reshapes the network organization that governs seizure susceptibility.

This study has limitations. Circadian concordance was established retrospectively in a limited-sized cohort of 38 participants with bilateral hippocampal RNS implants, and the generalizability of the biomarker to a larger and more diverse cohort of RNS implant configurations remains to be established. The closed-loop design of RNS precludes separability of stimulation from the interictal epileptiform activity (IEA) that triggers it. Therefore, it is possible that individuals whose IEA is more temporally concentrated over a day are intrinsically more responsive to treatment. Additionally, we did not account for effects of stimulation parameters or medication, which may independently influence treatment outcomes beyond the temporal patterning of stimulation examined here^4,44^. RNS ECoG snapshots are sampled intermittently, so sampling bias is possible, although novel application of sliding-window LS analysis provided a temporally coarse representation of long-term dynamics. Future work should explore Kalman filtering approaches which capture more fine-grained temporal dynamics by estimating the instantaneous phase of circadian cycles from longitudinal recordings^43,45^. Kuramoto modelling relied exclusively on the initial and final phase observations of each six-month epoch, restricting higher-resolution view of circadian phase trajectories and the use of more expressive oscillator models, such as the Stuart-Landau model^46^. Next generation devices may permit deliberate temporal patterning of stimulation and programming of cycle-based network biomarkers that would enable prospective validation of our *in silico* stimulation optimization pipeline for clinical translation.

Our study identifies circadian stimulation timing as a new dimension of therapeutic control for neurostimulation. Clinically, the *in silico* optimization framework offers a principled pathway towards personalized treatment and accelerated stimulation parameter tuning. This could advance contemporary practice, in which clinicians empirically navigate a vast stimulation parameter space without knowing whether or where more effective settings may exist. Hippocampal network circadian dynamics could also inform pre-implantation surgical planning and device placement by identifying recording or stimulation sites whose rhythms most closely track seizure risk. More broadly, these findings suggest that effective neurostimulation may require tuning not only where^47^, how strongly, or under what detection threshold stimulation is delivered, but also *when* it is delivered relative to the brain’s endogenous rhythms.

Key mechanistic questions remain for future investigation. First, does circadian concordance protect against seizures or reflect stimulation-induced network repair^29,48^? Second, do alpha band dynamics actively mediate the therapeutic effects of stimulation? Finally, how do circadian rhythms bridge the acute effects of stimulation with the chronic reorganization of brain networks, potentially through sleep-dependent plasticity? Although developed in epilepsy on the circadian timescale, our framework may generalize to other neurological and psychiatric disorders with rhythmic fluctuations on circadian and multidien timescales.

## Methods

### Participant cohorts and clinical outcomes

Study participants were individuals with drug-resistant epilepsy who had been implanted with bilateral hippocampal leads connected to a responsive neurostimulation (RNS) device (NeuroPace). Clinical outcomes were assessed longitudinally at scheduled clinic visits throughout the course of treatment. At each data-complete visit, clinical seizure frequency was assessed as the participant-reported number of seizures experienced in the preceding month. Seizure reduction at each visit was computed by comparing the fractional change in clinical seizure frequency compared to pre-implantation 3-month baseline, and was defined on a continuous scale from 0 (no reduction relative to baseline) to 1 (complete seizure freedom). In cases where the reported seizure rate exceeded the baseline rate, seizure reduction was assigned a value of 0 to reflect the absence of therapeutic benefit. Average seizure reduction over the full treatment course was computed as a weighted average of the longitudinal seizure reduction values, with weights proportional to the number of days elapsed between successive data-complete visits. Responder status was assigned both at the level of individual clinic visits and for each participant overall, based on average seizure reduction. Three categories were defined: super responder (seizure reduction > 0.9), intermediate responder (0.5 ≤ seizure reduction ≤ 0.9), and poor responder (seizure reduction < 0.5).

### Device data acquisition

Each participant was implanted with two intracranial leads, one in the left hippocampus and one in the right hippocampus. Each lead contained four electrode contacts arranged in a bipolar montage, yielding two bipolar channels per lead and four channels per participant in total. Channels 1 and 2 corresponded to the left hippocampus and Channels 3 and 4 to the right hippocampus.

The RNS device continuously acquired two types of data throughout the course of therapy. First, scheduled electrocorticography (ECoG) recordings were collected as 90-second clips at six-hour intervals, at times of day predetermined by the treating epileptologist for each participant (Fig. S1). The number and times of Scheduled ECoGs were generally evenly distributed within participants. These recordings were predominantly interictal and were acquired longitudinally across the full duration of device implantation. ECoG signals were sampled at 250 Hz. Second, the device recorded the cumulative number of stimulations delivered during each hour of the day, providing a continuous log of stimulation activity over the entire treatment course.

### ECoG data pre-processing and feature extraction

Each 90-second Scheduled ECoG clip was preprocessed independently through three sequential steps: bandpass filtering, stimulation artifact removal, and functional connectivity (FC) feature extraction. Signals were bandpass filtered into four physiologically defined frequency bands: theta (4–7 Hz), alpha (8–13 Hz), beta (14–30 Hz), and gamma (30–70 Hz). Filtering was performed using a second-order Butterworth filter applied in a forward-backward (zero-phase) fashion to prevent phase distortion. Notch filtering at 60 Hz was omitted, as the RNS device is battery-powered and does not introduce mains electrical interference. Stimulation artifacts were identified and removed prior to FC computation. For any stimulation event detected within a recording, the corresponding signal segment was excised along with a 1-second buffer on either side to eliminate residual artefactual activity. Excision was applied simultaneously across all channels at the affected timepoints to preserve cross-channel temporal alignment. Recordings in which residual artifact-free data totaled fewer than 30 seconds after excision were excluded from further analysis. Additional quality control criteria were applied to exclude recordings in which one or more channels lacked data, contained not-a-number (NaN) values, or exhibited temporal overlap with another scheduled recording.

FC was quantified using the phase locking value (PLV), a measure of inter-channel phase synchrony^12^. We used PLV as a measure of FC due to its robustness to interictal epileptiform activity (IEA)^14^. Instantaneous phase was extracted from each bandpass-filtered signal using the Hilbert transform and used to compute PLV between channel pairs. To characterize local hippocampal network dynamics, FC was computed exclusively between the two channels on the same lead, i.e., within the left hippocampus (Channels 1–2) and within the right hippocampus (Channels 3–4) independently. This yielded one PLV estimate per hemisphere per frequency band per recording, and two PLV estimates in total per band across both hippocampi. FC was extracted from all retained Scheduled recordings for each participant. This ensured consistent temporal alignment across recordings and facilitated downstream circadian analysis.

### Lomb-Scargle analysis of circadian cycles

Lomb-Scargle (LS) spectral analysis was used to characterize circadian rhythmicity in the sparse longitudinal FC timeseries^49,50^. Although scheduled ECoG recordings are acquired at nominally six-hour intervals, recordings are stored in the limited onboard memory of the RNS device and may be overwritten before being uploaded to the cloud by the participant. This results in a substantial proportion of missing timepoints, rendering the FC timeseries effectively irregularly sampled. Conventional spectral methods assume uniform sampling and are therefore unsuitable for this setting^51^. LS analysis circumvents this limitation by performing regression against sinusoidal basis functions directly on the observed, unevenly spaced data.

Circadian dynamics were quantified by fitting an LS model with a fixed 24-hour period to the FC timeseries. Specifically, the model regressed the observed PLV values against sine and cosine basis functions with period *T* = 24 hours by solving the following optimization:

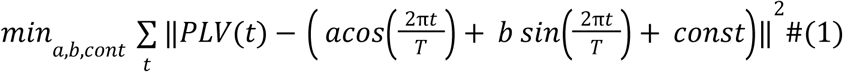

From these coefficients, the amplitude (*A^c^*) and peak phase (*Φ^c^*) of the circadian FC cycle were derived as:

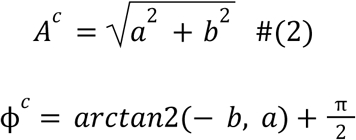

LS models were fitted independently for each frequency band and each FC channel pair (hemisphere). To track changes in circadian FC dynamics over time, LS models were fitted within non-overlapping six-month windows (epochs) anchored to the date of device implantation. There were two challenges with the application of LS analysis in this study. First, because circadian peak phase can shift within a six-month window, the superposition of phase-shifted cycles across recordings may attenuate the apparent cycle amplitude, potentially leading to underestimation of true circadian rhythmicity. Second, while biological circadian periods naturally jitter around 24 hours, the LS model fitted here assumes a fixed period of exactly 24 hours, which may introduce additional underestimation of cycle strength when the true period deviates from this value. To mitigate these sources of error, analyses were restricted to six-month windows containing at least 120 scheduled recordings, ensuring sufficient sampling density for reliable spectral estimation. Participants who did not contribute at least one qualifying six-month window were excluded from the study. This criterion resulted in a final cohort of 38 participants (Supplementary Table S1). LS cycles at each six-month window were time-matched to a subsequent clinic visit within the following 1 year. If multiple six-month intervals were matched to the same clinic visit, then only the most recent one was considered as time-matched.

### Measures of network circadian cycling

Two summary measures of hippocampal network circadian dynamics were derived from the LS-estimated cycle amplitudes and peak hours for each frequency band and six-month epoch.

*Circadian amplitude* (CA) quantified the overall strength of circadian cycling in the hippocampal network. CA was computed as the sum of the LS-estimated amplitudes of the left (*A^l^*) and right (*A^r^*) hippocampal FC cycles:

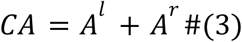

*Circadian concordance* (CC) quantified the degree of bilateral alignment in the circadian timing of hippocampal network activity. CC was defined as one minus the circular variance of the left (Φ*^l^*) and right (Φ*^r^*) hippocampal circadian peak phases:

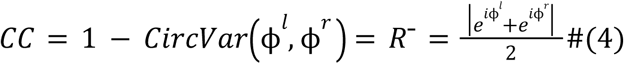

where *R*^-^ is the mean resultant length of the corresponding unit vectors on the unit circle. CC ranges from 0, indicating maximal phase dispersion (peak hours 12 hours apart), to 1, indicating perfect bilateral synchrony (identical peak hours). Both CA and CC were computed separately for each frequency band and six-month epoch.

### Stimulation data preprocessing

Stimulation activity was characterized using the continuous hourly stimulation count log recorded by the RNS device. For each participant, hourly stimulation counts were averaged across all available days for each hour of the day (0–23), yielding a 24-point profile of mean stimulation activity. This profile was then min-max normalized across the 24-hour period within each participant, mapping the minimum mean hourly count to 0 and the maximum to 1, with all intermediate values scaled linearly. This normalization reflects the assumption that neural tissue adapts to the prevailing level of stimulation, such that the minimum delivered stimulation approaches a quiescent baseline, maximum stimulation approaches saturation, and intermediate values scale linearly. All downstream analyses were performed on this normalized average stimulation profile. For longitudinal analyses, the same averaging and normalization procedure was applied within non-overlapping six-month windows anchored to the date of implantation, yielding a time-resolved stimulation profile for each epoch.

Two scalar features were derived from the normalized stimulation profile for each interval of interest. The *peak hour of stimulation* was defined as the hour of the day at which the normalized average stimulation reached its maximum value (i.e., equal to 1). The *stimulation profile area* (SPA) was defined as the sum of the normalized average stimulation values over a specified set of hours, quantifying the temporal concentration of stimulation within that interval. SPA was computed for two interval definitions: the full 24-hour period (SPA(24h)), and a specified sub-interval between hours h1 and h2 (SPA(h1–h2)). Higher SPA values indicate stimulation that remains elevated over a broader portion of the day.

### Multivariate regression and ML modeling with recursive feature elimination

Multivariate linear regression was employed for four distinct analyses: (i) biomarker discovery, relating network circadian features to seizure reduction; (ii) prediction of longitudinal seizure reduction from circadian and stimulation features; (iii) identification of factors driving changes in circadian peak hour over successive six-month epochs; and (iv) prediction of Kuramoto model coupling constants from circadian cycle and stimulation features. All regression models were implemented using the Python statsmodels library^52^.

For each regression target, an initial candidate feature set was assembled and subjected to iterative recursive feature elimination (RFE) within a leave-one-participant-out (LOPO) cross-validation framework. At each iteration, the model was trained across all cross-validation folds and the number of folds in which each feature was a statistically significant predictor of the dependent variable was recorded. Features that were significant in at least one fold were retained; all others were discarded. This reduced feature set was then used as the input to the next RFE iteration. The process continued until either no features remained (indicating an absence of reliable predictive signal) or all remaining features met the significance criterion in at least one fold. Feature significance was assessed using the training data within each fold to prevent information leakage from the test set. Prior to model fitting within each fold, all features were z-scored using the mean and standard deviation computed from the training data only, preventing any leakage of test-set distributional information into the normalization step.

LOPO cross-validation was adopted throughout to provide a conservative, clinically relevant estimate of model generalization and to avoid overestimation of effect sizes. Because some participants contributed very few observations (in some cases a single sample), fold-level mean squared error (MSE) estimates were highly variable and unsuitable for direct averaging. To address this, test-set predictions from all folds were pooled across participants and compared jointly against the corresponding ground-truth values. Predictive performance was quantified as the Pearson correlation (*r*) between pooled predictions and ground truth. Model MSE was similarly computed on the pooled predictions. A baseline model was defined as the participant-agnostic mean predictor, in which the mean of the dependent variable computed from the training data in each fold was used as the prediction for all test observations in that fold. Baseline MSE was computed from the corresponding pooled baseline predictions to ensure a directly comparable reference. Percentage improvement in MSE over baseline was used to report the relative gain achieved by each model.

Logistic regression was employed for binary classification tasks, with feature selection conducted using the same iterative RFE framework described above. At each RFE iteration, features were retained if their unit-normalized absolute regression coefficient exceeded 0.05 in at least one LOPO cross-validation fold, providing a consistent threshold for identifying features with non-negligible discriminative contribution. All features were z-scored using the mean and standard deviation computed from the training data only prior to model fitting within each fold. LOPO cross-validation was applied both during RFE and for final performance evaluation. As with the regression models, test-set predictions were pooled across all folds, and the area under the receiver operating characteristic curve (AUROC) was used as the primary performance metric.

### Kuramoto model description and training

The Kuramoto model describes the temporal evolution of phase in a system of coupled oscillators and was used here to characterize how hippocampal network circadian peak phases change over time under the influence of stimulation and inter-regional interactions. The model comprised five oscillators, each with a fixed period of 24-hours, corresponding to: (i) the left hippocampal FC circadian cycle, (ii) the right hippocampal FC circadian cycle, (iii) the circadian profile of stimulation delivery, (iv) a diurnal attractor with a fixed peak phase at 3 PM, and (v) a nocturnal attractor with a fixed peak phase at 3 AM. Instantaneous phases of these oscillators are 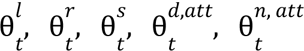 respectively. The attractor peak phases were set based on the empirically observed cluster centres in the population-level polar histogram of circadian peak hours and approximated to 3AM and 3PM for computational ease (Fig. 3a). The phase dynamics of each oscillator were governed by the Kuramoto equations:

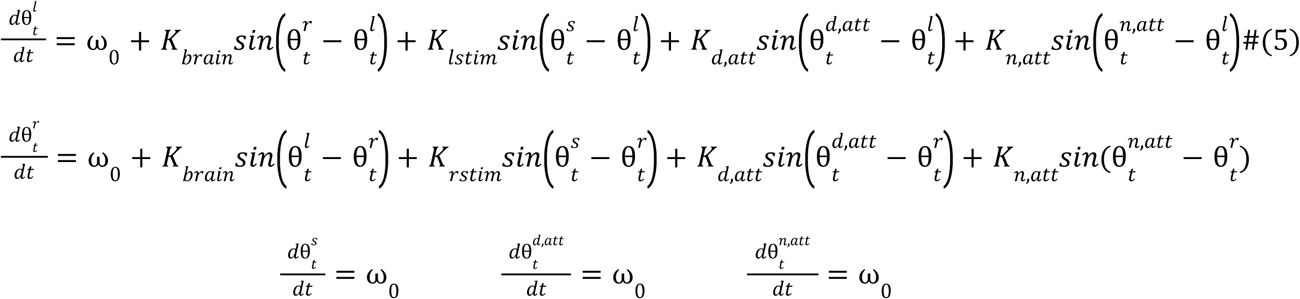

Five coupling constants parameterized the inter-oscillator interactions. *K_brain_* captured the mutual coupling between the left and right hippocampal FC circadian cycles. *K_lstim_* and *K_rstim_* captured the influence of stimulation on the left and right network cycles, respectively. *K_d_*_,*att*_ and *K_n_*_,*att*_ captured the influence of the diurnal and nocturnal attractor oscillators, respectively. We assumed that for each network cycle, only the attractor whose peak phase was closest to the current cycle peak phase exerted an influence, thereby implementing a winner-take-all attractor mechanism consistent with the two-cluster structure observed in the data. This allowed the effect of the attractors to be summarized by a single coupling constant *K_att_* corresponding to the attractor that was closer to the network cycle: 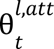 and 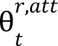 for the left and right cycle, respectively. The stimulation oscillator and both attractor oscillators were treated as external drivers; they were not influenced by any other oscillator in the system, and their peak phases therefore remained fixed throughout the simulation. Under these assumptions, the equations of the network oscillators simplify, each expressing the rate of change of a network cycle’s phase as a function of three influences: (i) the contralateral hippocampal network cycle, (ii) stimulation, and (iii) the nearest attractor:

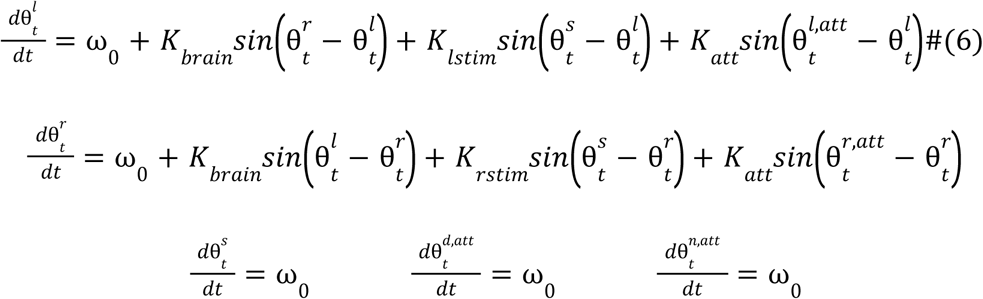

Preliminary analyses revealed that coupling constants varied substantially across participants and across epochs within the same participant. Coupling constants were therefore estimated independently for each six-month epoch. Each epoch was defined by a set 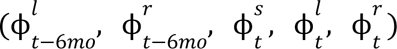, comprising the circadian cycle features (peak phase of the left and right hippocampal cycles) at the start of the window (*t* − 6 months) and at the end of the window (*t*), together with the stimulation characteristics during the window.

Coupling constants were estimated from the observed terminal peak phases of the network cycles, the stimulation oscillator, and the attractors using a custom optimization algorithm. Conventional Kuramoto parameter estimation requires a dense timeseries of phase observations from the initial state to final timepoint^53,54^. Since only the initial and final peak phases were available for each six-month epoch in our data, we developed a steady-state approximation approach. We assumed that over the course of 6 months, the system reached a new stable phase configuration relative to the other oscillators in the system, and that the peak phases measured at the end of the window 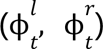 represent this converged state. Under the steady-state assumption, the relative phase between oscillators no longer changes. Given that all oscillators share the same period (eliminating frequency-driven phase drift), the differential equations simplify to a static system in which instantaneous phase differences can be substituted by the differences between peak phases:

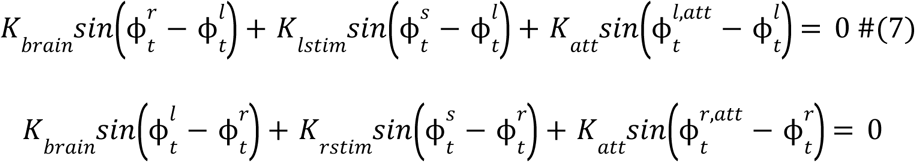

This system is under-determined, yielding two equations in four unknowns *K_brain_*, *K_lstim_*, *K_rstim_*, *K_att_.* To obtain a unique solution, two constraints were imposed. First, *K_att_* was fixed at 2 without loss of generality, as its sign (positive) is determined by its role as an attractor and its magnitude sets the scale for the remaining coupling constants. Second, the minimum-norm least-squares solution was selected among all solutions satisfying the linear system, providing a unique and well-defined estimate of the three free coupling constants. This procedure was repeated for each epoch to obtain sample-specific estimates of *K_brain_*, *K_lstim_*, *K_rstim_*.

The quality of the estimated coupling constants was assessed by evaluating their ability to reproduce the empirically observed circadian peak hours at the end of each six-month epoch. For a given epoch, the estimated coupling constants were used together with the initial peak phases 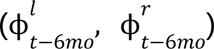 and stimulation peak phase 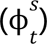 to initialise the Kuramoto model, which was then integrated forward for 180 days using a step size of 0.001d. The predicted peak hours at 180 days were compared against the observed terminal peak phases 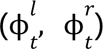. Prediction error was defined as the sum of the absolute hourly differences between predicted and observed peak hours for the left and right hemispheres. Only epochs for which the forward simulation had converged were retained for evaluation, with convergence measured as the sum of magnitude of the LHS in the two equations in Eq (7) being below 1e-5.

We note that this validation scheme estimates and evaluates coupling constants on the same set, which departs from conventional train-test separation in machine learning studies, but is not uncommon in Kuramoto literature. We argue that this approach is nonetheless non-trivial and appropriate for four reasons. First, coupling constants are sample-specific by design, as they vary across epochs and participants; a held-out evaluation scheme is therefore not applicable in the conventional sense. Second, the Kuramoto equations are non-linear, such that even small perturbations in coupling constants can produce qualitatively different phase trajectories; achieving low prediction error thus constitutes a meaningful constraint on the solution. Third, using the entire timeseries for parameter estimation is a standard procedure in several previous applications of Kuramoto models to empirical neurophysiological data^53,54^. Fourth, the coupling constants were obtained from an under-determined system subject to a minimum-norm constraint; achieving low prediction error under these conditions is non-trivial, as it is not guaranteed that biologically arbitrary constraints would yield accurate forward simulations. We further acknowledge that the steady-state approximation does not account for the initial phase configuration of the system, which is known to influence trajectory dynamics in non-linear systems. These are inherent limitations of having access only to the start- and end-point phases of each epoch, rather than a continuous phase timeseries. These limitations should be interpreted as estimation and data limitations rather than fundamental incompatibilities between the Kuramoto framework and the underlying biology.

The full Kuramoto model was compared against two baseline models, both sharing the same non-linear differential equation structure to enable a controlled assessment of the incremental contribution of each coupling term. Baseline 1 retained only the attractor coupling, with *K_att_* = 2 *and K_brain_* = *K_lstim_* = *K_rstim_* = 0. This model captured only the variance in long-term peak phase dynamics attributable to the diurnal and nocturnal attractors, without any inter-hemispheric or stimulation-driven coupling. Baseline 2 additionally included inter-hemispheric coupling, with *K_att_* = 15*, K_brain_* estimated via least-squares, and *K_lstim_*= *K_rstim_* = 0. This model allowed attractor dynamics and inter-regional synchronisation to jointly explain circadian peak phase evolution, while excluding any direct effect of stimulation. Both baseline models were fitted and evaluated using an identical procedure applied to the full model.

### In silico optimization of stimulation using Kuramoto model

The validated Kuramoto framework was used to perform *in silico* optimization of stimulation delivery. For a given participant’s initial circadian cycle configuration, we asked: what peak hour of stimulation would maximize bilateral alignment of hippocampal circadian cycles over a six-month horizon? Circadian concordance was used as the optimization target, given its established role as a biomarker of ongoing seizure reduction. The peak hour of stimulation was treated as the free parameter, consistent with our empirical finding that stimulation timing influences the long-term dynamics of circadian peak phases.

For each initial configuration of left and right hippocampal circadian cycle peak phases, the Kuramoto model was run forward for 180 days for each of the 24 candidate stimulation peak hours (midnight through 11 PM), in one-hour increments. For each candidate hour, the predicted terminal peak phases of the left and right hippocampal cycles were used to compute circadian concordance as an approximation of the expected seizure reduction under that stimulation setting. The stimulation peak hour yielding the highest circadian concordance was identified as the optimal setting for that initial configuration. To demonstrate the clinical applicability of this approach, all *in silico* analyses were conducted in a strictly held-out fashion. For each participant, coupling constant prediction models (described below) were evaluated using only the fold in which that participant served as the test set, ensuring that no data from the participant under analysis contributed to any aspect of model training.

The outcome of forward Kuramoto simulation depends on the coupling constants *K_brain_*, *K_lstim_*, *K_rstim_*. Since these constants must be specified prospectively for a new six-month epoch, i.e., before the terminal cycle state is observed, predictive models were developed to estimate them from features available at the start of the epoch: the initial circadian cycle features 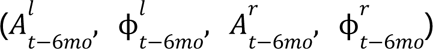 and stimulation characteristics (peak hour 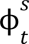 and derived SPA measures). For prospective prediction in a new six-month epoch, the coupling constants for a given participant were predicted using only the model trained on other participants (i.e., the corresponding held-out LOPO fold), ensuring no within-participant data leakage.

Ground-truth coupling constants for model training were obtained from the estimates derived from real data as described in the Kuramoto model fitting section. Circadian cycle amplitudes and SPA measures were included as additional predictors alongside the initial peak phase features. *K_brain_* was predicted using a multivariate linear regression model with RFE, trained and evaluated under LOPO cross-validation. To predict stimulation–cycle coupling strengths *K_lstim_*, *K_rstim_* were modelled jointly using a pooled hierarchical regression model, combining both left and right coupling estimates into a single training set. A hierarchical modelling approach was adopted based on preliminary analyses. Model performance was evaluated using LOPO cross-validation by comparing predicted coupling strengths against the empirically estimated values from real data. Prior to forward simulation, LOPO predicted coupling strengths were scaled to match the scale of the empirical coupling values.

We estimated SPA from a given candidate stimulation peak hour for two reasons: (i) SPA features were important predictors of coupling constants, and (ii) SPA is not directly observable for a hypothetical stimulation peak hour during *in silico* analysis. SPA(7AM–11AM) was found to be well approximated as a sinusoidal function of stimulation peak hour in preliminary analyses, and a sinusoidal model was fitted to the observed data for this purpose (Fig. S10). For SPA(11AM–7AM), no consistent functional relationship with stimulation peak hour was identified. Instead, for each candidate peak hour, the median of the SPA(11AM–7AM) values observed across participants at that stimulation peak hour in the real data was used as the estimate. For candidate peak hours at which no participant’s stimulation had been observed to peak, the mean of the hour-level medians across all observed hours was substituted as a fallback estimate.

### Hierarchical regression model

Preliminary analyses revealed that the relationship between circadian cycle amplitude and stimulation–cycle coupling constants (*K_lstim_*, *K_rstim_*) was non-linear. Initial cycle amplitude 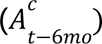 influenced the magnitude of the coupling but coupling itself could be positive or negative. Thus, a two-stage hierarchical regression model was developed in which the sign of the amplitude was treated as a latent variable to be predicted, yielding an effective signed amplitude feature that entered the final regression. The model takes the form:

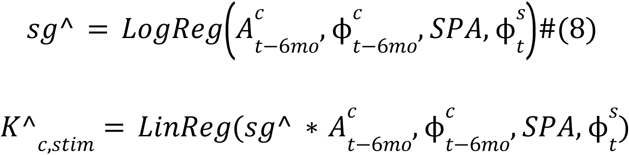

The function adjusts the sign of the amplitude contribution before it enters the linear regression, introducing a sample-specific non-linearity into an otherwise linear model. The model was trained in two sequential stages. In the first stage, a logistic regression classifier was trained to predict the sign *sg* ^ of the coupling in each sample. Feature selection was performed using RFE with LOPO cross-validation. Model fitting in this stage was performed exclusively on the training data within the cross-validation fold. In the second stage, a multivariate linear regression model was trained to predict the stimulation–cycle coupling constant. The feature set included the signed amplitude term 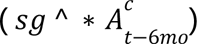 that was computed using the sign predicted by the first-stage classifier alongside other candidate features subjected to RFE.

For prediction on a held-out test sample, the two stages were applied sequentially: the first-stage logistic model, trained on all participants except the test participant, was used to predict the sign *sg* ^ for the test sample. The resulting signed amplitude, together with the other RFE-selected features, was then passed to the second-stage linear model to generate the final coupling constant prediction. Model performance was assessed by pooling test-set predictions across all LOPO cross-validation folds and comparing them against the empirical coupling constants estimated from the Kuramoto model fitting procedure.

## Data Availability

Raw RNS System data were obtained through a Data Use Agreement between UCSF and NeuroPace and therefore cannot be publicly shared.

## Acknowledgements

This work was supported by Schmidt Science Fellows, CURE: Taking Flight Award, and the Chen Scholars Program at UCSF, funded by Chrissy Luo and Tianqiao Chen.

## Supplementary Information: Cycle-based neurostimulation for control of circadian brain network dynamics in human epilepsy

**Table S1.** Participant Demographics.

| # | ID | Sex | †Age at Epilepsy Onset | Epilepsy Etiology | MRI findings (hippocampi) | †Age at RNS Implant | RNS Data Duration (days) | Scheduled iEEG Recordings (#) | Average seizure reduction |
|---|---|---|---|---|---|---|---|---|---|
| 1 | NP1 | M | 10–19 | unknown | normal | 30–39 | 3715 | 2917 | 0.87 |
| 2 | NP5 | M | 10–19 | unknown | left HS | 40–49 | 3383 | 11468 | 0.81 |
| 3 | NP15 | M | < 10 | TBI | bilateral HS | 40–49 | 2062 | 699 | 0.67 |
| 4 | NP17 | F | 10–19 | ecclampsia | bilateral HS | 30–39 | 2108 | 1754 | 0.56 |
| 5 | NP19 | F | 10–19 | unknown | normal | 20–29 | 2589 | 1399 | 0.97 |
| 6 | NP21 | F | < 10 | unknown | normal | 60–69 | 2753 | 2003 | 0.94 |
| 7 | NP29 | M | unknown | unknown | unknown | 60-69 | 2648 | 9512 | 1 |
| 9 | NP34 | M | 20–29 | unknown | left HS | 40–49 | 2539 | 1590 | 0.76 |
| 10 | NP36 | M | 30–39 | encephalitis | bilateral HS | 30–39 | 2550 | 5132 | 0.99 |
| 11 | NP38 | M | 40–49 | unknown | normal | 50–59 | 2529 | 1320 | 0.97 |
| 12 | NP39 | F | 10–19 | encephalitis | normal | 20–29 | 2424 | 6009 | 0.09 |
| 13 | NP41 | M | 20–29 | unknown | normal | 20-29 | 512 | 447 | 0.61 |
| 14 | NP43 | F | < 10 | unknown | bilateral HS | 30–39 | 2459 | 764 | 0.56 |
| 15 | NP45 | F | 30–39 | epidermal nevus syndrome | normal | 40–49 | 2363 | 761 | 0.54 |
| 16 | NP47 | M | < 10 | meningitis | normal | 20–29 | 1661 | 1978 | 1 |
| 17 | NP52 | F | 30-39 | encephalitis | bilateral HS | 30-39 | 2074 | 2280 | 0.69 |
| 18 | NP55 | M | 30–39 | TBI | normal | 40–49 | 2556 | 3655 | 0.59 |
| 19 | NP56 | M | 30-39 | unknown | unknown | 30-39 | 1812 | 1112 | 0.54 |
| 20 | NP61 | F | unknown | unknown | unknown | 20-29 | 2201 | 1423 | 0.8 |
| 21 | NP62 | F | 40-49 | meningoencephalocele | bilateral HS | 50-59 | 1885 | 3257 | 0.74 |
| 22 | NP63 | M | < 10 | meningoencephalocele | right HS | 30-40 | 1864 | 1998 | 0.89 |
| 23 | NP68 | M | 30-39 | TBI | bilateral HS | 60-69 | 1770 | 5053 | 0.8 |
| 24 | NP70 | F | 20-29 | unknown | bilateral HS | 30-39 | 1167 | 1887 | 0.5 |
| 25 | NP72 | F | 20-29 | unknown | bilateral HS | 30-39 | 1640 | 656 | 0.21 |
| 26 | NP86 | M | < 10 | unknown | normal | 30-39 | 1436 | 1794 | 0 |
| 27 | NP87 | M | 60-69 | unknown | left HS | 60-69 | 1400 | 5263 | 0.21 |
| 28 | NP88 | M | 20-29 | unknown | left HS | 30-39 | 1380 | 2173 | 0.67 |
| 29 | NP91 | F | 20-29 | unknown | normal | 30-39 | 1267 | 3754 | 1 |
| 30 | NP95 | F | 10-19 | unknown | normal | 30-39 | 1240 | 3084 | 0.29 |
| 31 | NP96 | M | 20-29 | unknown | left HS | 30-39 | 1233 | 2551 | 0.54 |
| 32 | NP99 | M | unknown | unknown | unknown | 20-29 | 1192 | 3016 | 0 |
| 33 | NP102 | M | 30-39 | unknown | normal | 40-49 | 911 | 2031 | 0.15 |
| 34 | NP103 | F | 10-19 | unknown | normal | 20-29 | 904 | 1050 | 0.2 |
| 35 | NP104 | F | < 10 | unknown | normal | 10-19 | 881 | 2956 | 0.22 |
| 36 | NP120 | F | 20-29 | unknown | bilateral HS | 30-39 | 126 | 418 | 0.86 |
| 37 | NP123 | M | 20-29 | TBI | bilateral HS | 20-29 | 414 | 555 | 0.85 |
| 38 | NP125 | F | 20-29 | unknown | bilateral HS | 20-29 | 372 | 1175 | 0 |
| 39 | NP140 | M | < 10 | unknown | normal | 20-29 | 712 | 455 | 0.99 |

All participants (N=38) had Responsive Neurostimulation (RNS) System depth leads in the bilateral hippocampi
†Ages shown in ranges to minimize identifiable information
M, male; F, female; TBI, traumatic brain injury; MRI, magnetic resonance imaging;

### Participant clinic visits and device programming

Patients undergoing RNS therapy typically receive clinical follow-up approximately every three months, during which monthly clinical seizure frequency is assessed. In this study, we restricted our investigation to data-complete visits for which outcomes data were available for retrospective analysis. Across 38 participants, we analyzed 145 data-complete visits (median 3 per participant, IQR 2–5) with median interval of 259 days (IQR 140–628 days).

**Figure S1:**
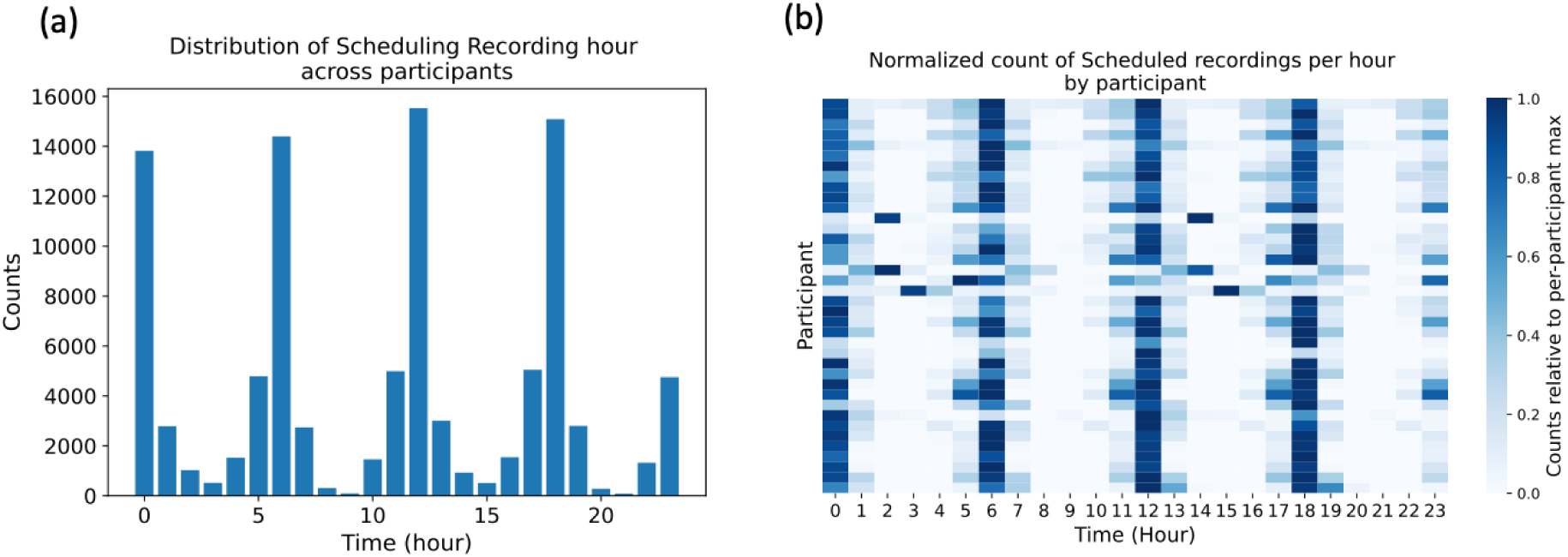
Timing of ECoG Scheduled recordings. (a) Hourly distribution of scheduled recording trigger time aggregated across the entire data of all the participants. (b) Hourly distribution of scheduled recording trigger time within each participant, normalized to the participant-specific maximum count. In many participants, recordings were scheduled for every 6 hours, accounting for the modes of timings.

### Characterization of network circadian cycles

**Figure S2:**
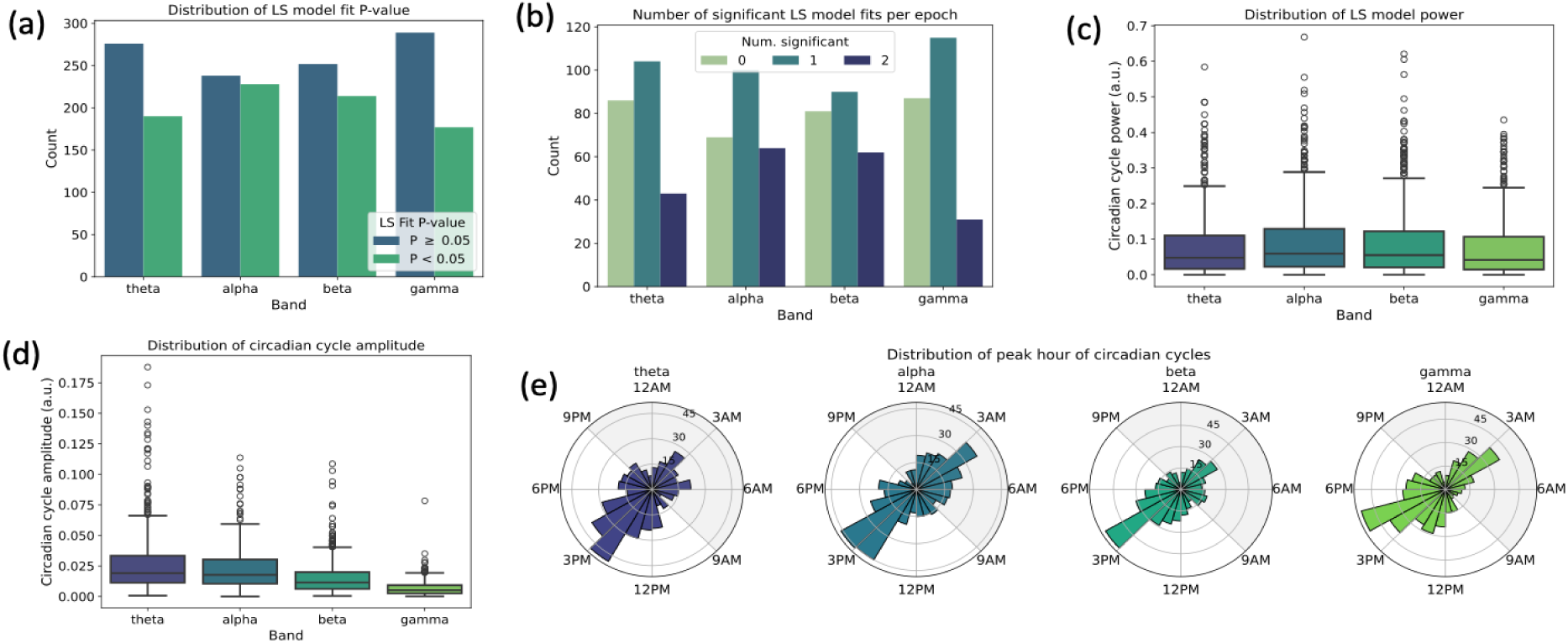
Lomb-Scargle model fit assessment and circadian cycle characteristics. (a) Band-wise distribution of P-values of Lomb-Scargle model fit across participants, hemispheres, and epochs. Network circadian cycle prevalence was frequency-dependent, being most abundant in the alpha band (*n* = 228/466, 48.9% with significant LS model fits) and least abundant in the gamma band (*n* = 177/466, 38.0%). To avoid selection bias, all epochs were retained in the analyses regardless of LS fit significance; the robustness of this choice was assessed in a sensitivity analysis. (b) Band-wise distribution of number of significant model fits in each six-month epoch across participants. Max per epoch is two when LS fit for both left and right hippocampus is significant. (c, d, e) Band-wise distribution of circadian cycle power, amplitude, and peak hour across hemispheres and six-month epochs from all the participants.

### Network circadian cycles concordance biomarker

**Figure S3:**
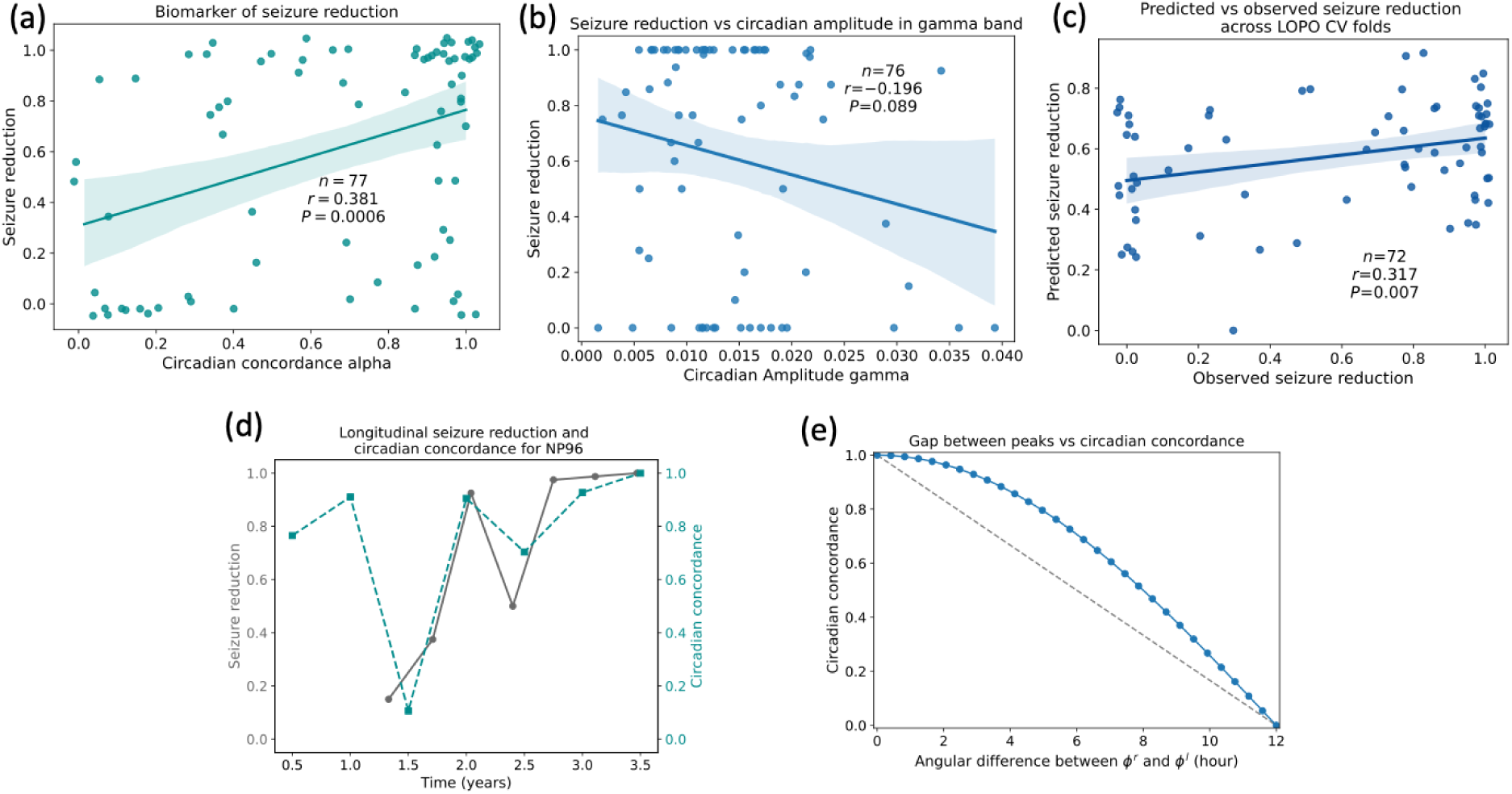
Circadian cycle biomarker. (a) Circadian concordance alpha is associated with time-matched longitudinal seizure reduction. Random jitter of 0.05 was added to the data for visualization purpose only. (b) Weak non-significant association between circadian amplitude in gamma band with time-matched longitudinal outcomes across participants. (c) Cross-validated model predictions for seizure reduction based on time-matched circadian features, along with log of baseline seizure rate. Random jitter of 0.03 was added to x-axis for visualization only. (d) Seizure reduction and circadian concordance in alpha band for an example participant. (e) Relationship between circadian concordance and angular difference of two cycles with peak phases at 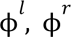.

### Robustness analysis of circadian cycles as biomarker of seizure reduction

To investigate the robustness of the circadian concordance biomarker, we re-analyzed the data with a stricter inclusion criterion. Only epochs in which at least one of the two alpha band network circadian cycles had a significant LS model fit were retained for the analysis.

With the stricter inclusion criterion, circadian concordance of the alpha band was significantly associated with ongoing seizure reduction (*n* = 56, *r* = 0.503, *P* < 0.001; Fig. S4a). Multivariate models that predicted outcome from circadian features achieved comparable or higher performance when epochs with significant LS model fit cycles were considered, compared to the case when all epochs were used (Fig. S4b). Furthermore, the peak phase of circadian cycles in alpha showed a bimodal distribution even when only significant LS model fit cycles were considered (Fig. S4c) (Kuiper test, *n* = 228, *P* = 3.2e-9; cluster centers: 3:18AM and 2:44PM; cluster boundaries: 8:37AM and 8:57PM). Overall, these results support the broader validity of the observation that alignment of network circadian cycles is a biomarker of ongoing seizure reduction, with the bimodal distribution of cycle peaks underpinning the biomarker.

**Figure S4:**
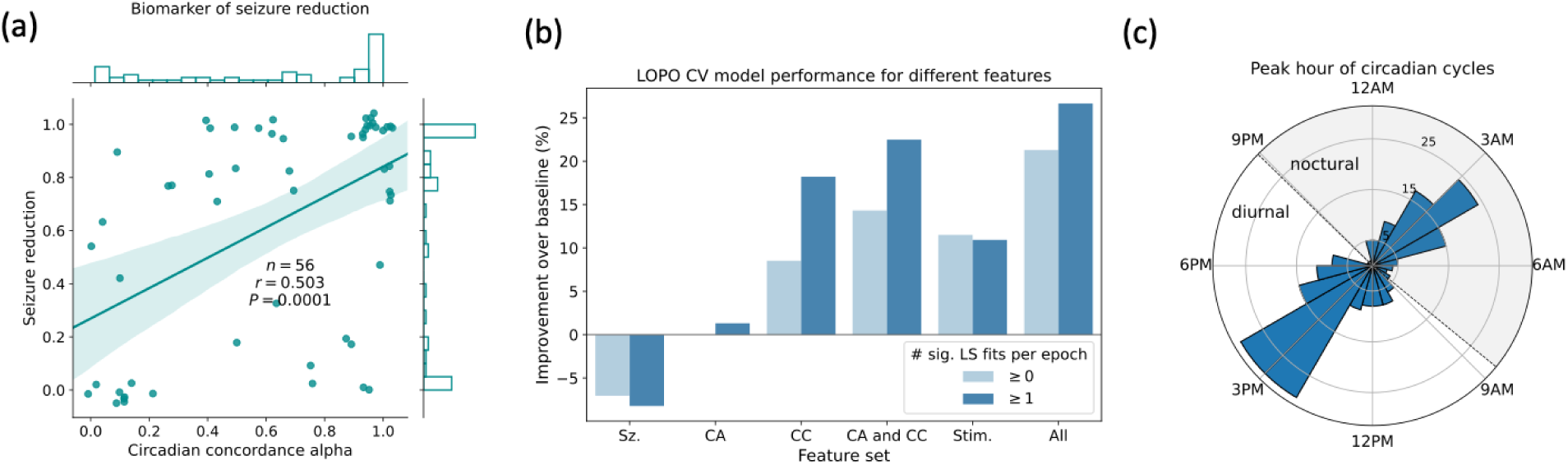
Biomarker discovery, outcome prediction, and peak clustering holds even with stricter inclusion criterion for cycles. (a) Circadian concordance in alpha band is associated with ongoing seizure reduction for epochs with at least one hippocampus with significant LS fit. Random jitter of 0.05 was added to the data for visualization purpose only; statistics and histogram were computed on unjittered values. (b) Outcome prediction using circadian features. All epochs (same as Fig. 2d and Fig. 4f) are compared to epochs in which at least one hippocampal LS model fit was significant. Improvement in prediction over population mean baseline is shown. (c) Distribution of peak hour of circadian cycle alpha with significant LS model fit across all participants and six-month epochs (*n* = 228). Peaks clustered around diurnal and nocturnal attractors. Boundaries between the clusters are shown in dotted lines.

### Circadian cycles of power- in-band features

We evaluated how cycling of spectral power-in-band (PIB) compares to cycling of functional connectivity (FC) as a predictor of ongoing seizure reduction. Data was pre-processed using the same pipeline as for the FC values. A single PIB value was computed for each channel and band from a Scheduled recording. Recordings were divided into non-overlapping six-month windows (epochs) from the date of implantation for each participant. Cycle characteristics were measured by fitting Lomb-Scargle models independently to each epoch and band across participants. Only epochs with at least 120 Scheduled recordings were included for downstream analysis.

Similar to network circadian cycles, PIB circadian cycles were prevalent across hippocampi (channels), bands, and participants. PIB circadian cycles were more prevalent than FC circadian cycles (Fig. S5). Cycle amplitudes were largest for theta band and smallest for gamma band (Fig. S5a). Cycles in theta, alpha, and beta bands showed a unimodal distribution with a peak around 3AM (Fig. S5b), which may reflect the sleep-wake cycle. The unimodal distribution of peaks in the alpha band was in contrast to the bimodal distribution of peaks from network circadian cycles. We explored the potential of PIB circadian cycles as a biomarker of ongoing seizure reduction. PIB circadian cycle features predicted seizure reduction better than population mean baseline (Fig. S5e). However, network circadian concordance were better predictors of outcome than power-based circadian concordance. When combined with stimulation features, network circadian cycles predicted outcomes better than power-based circadian cycles. Thus, circadian cycles are widely prevalent in ECoG data with network circadian cycles being most predictive of ongoing seizure reduction.

**Figure S5:**
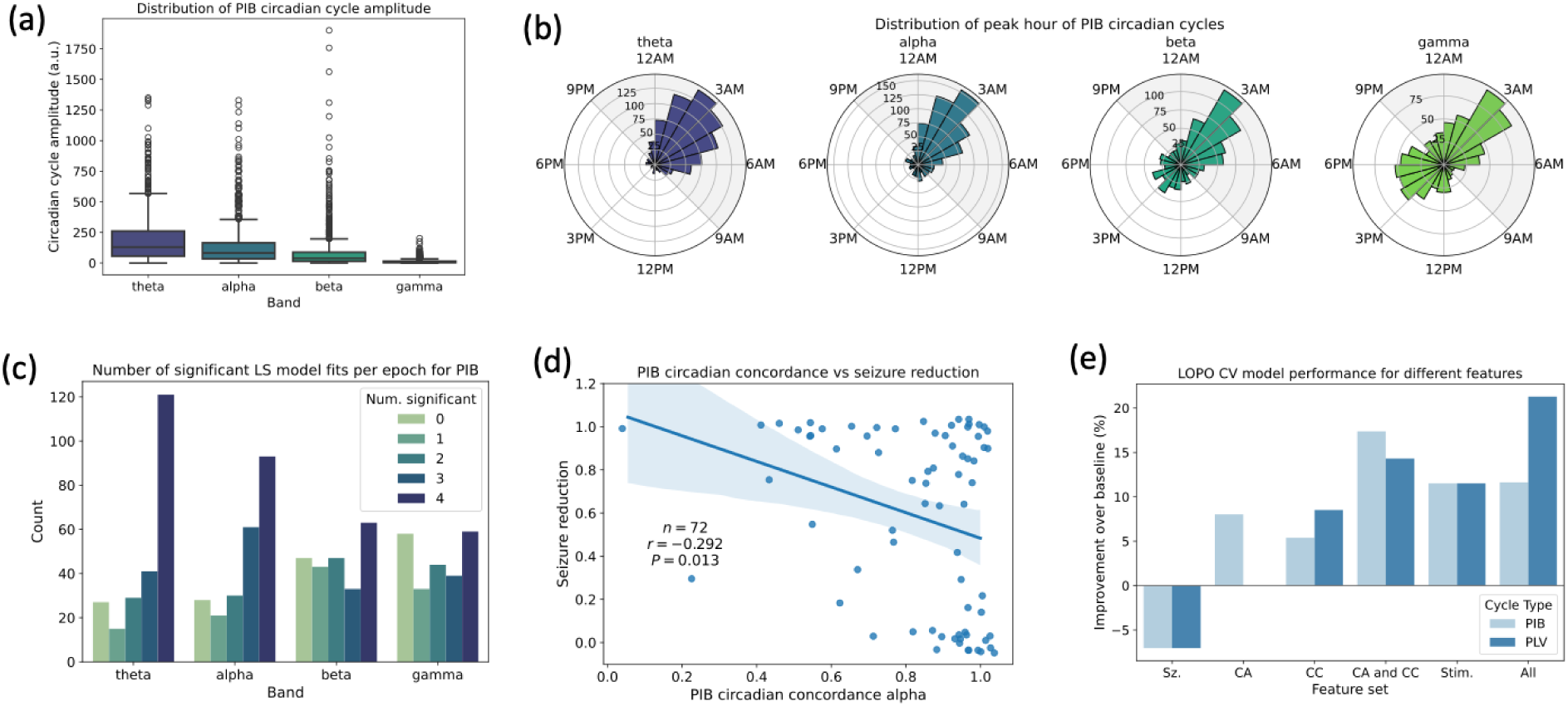
Power-in-Band (PIB) Circadian Cycles. (a, b) Band-wise distribution of amplitude and peak hour of circadian cycles of PIB values across channels and six-month epochs from all the participants. (c) Band-wise distribution of number of significant model fits in each six-month epoch across participants. Max per epoch is four when LS fit is significant for both channels in left and right hippocampus. (d) PIB circadian concordance in alpha band is associated with time-matched longitudinal seizure reduction. Random jitter of 0.05 was added to the data for visualization purpose only; statistics were computed on unjittered values. (e) Outcome prediction using circadian cycle features of PIB values compared to functional connectivity (PLV) values.

### Clusters of network circadian cycle peak hours

We performed Gaussian mixture model (GMM) clustering of the circadian cycle peaks to identify the noctural and diurnal attractors, i.e., cluster centers. Since the peak hour is circular, to enable clustering, we linearized the cycle with peak hour ℎ as a tuple 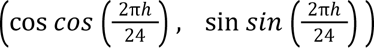. The linearized values were clustered into two clusters using GMM since the polar histogram of circadian cycle peaks showed two dominant clusters. The cluster mean and cluster boundaries were obtained from the fitted GMM. To find the cluster boundaries, we found the angle (time of day) at which the posterior probability of the phase belonging to either cluster was close to 0.5. GMM initialization and clustering was performed 100 times, and the cluster means and boundaries were averaged across the iterations. The cluster centers were 3:39AM ± 4.1mins for the noctural and 3:02PM ± 3.5mins for the diurnal cluster. Boundaries between clusters were at 9:29AM ± 13.5 mins and 8:57PM ± 3.1 mins. To simplify Kuramoto modeling, we approximated the cluster centers to be 3AM for the nocturnal cluster and 3PM for the diurnal cluster, and the boundaries to be at 9AM and 9PM.

### Change in circadian peak over six-month intervals

We studied whether inter-cluster transitions reflect a gradual drift in peak hour or an abrupt shift occurring within a single six-month window. To distinguish between these possibilities, we compared the magnitude of angular change in peak hour between successive windows for within-cluster and inter-cluster transitions (Fig. S6). On average, inter-cluster transitions were associated with a change of 5.8 ± 3.0 hours in peak hour, compared to 1.7 ± 1.7 hours for within-cluster transitions, indicating that transitions between clusters result from large, discrete shifts in circadian phase rather than slow monotonic drift. Importantly, within-cluster transitions were not without variability; the peak hour continued to fluctuate even in the absence of a state change, underscoring the dynamic nature of hippocampal network circadian organization across time.

**Figure S6:**
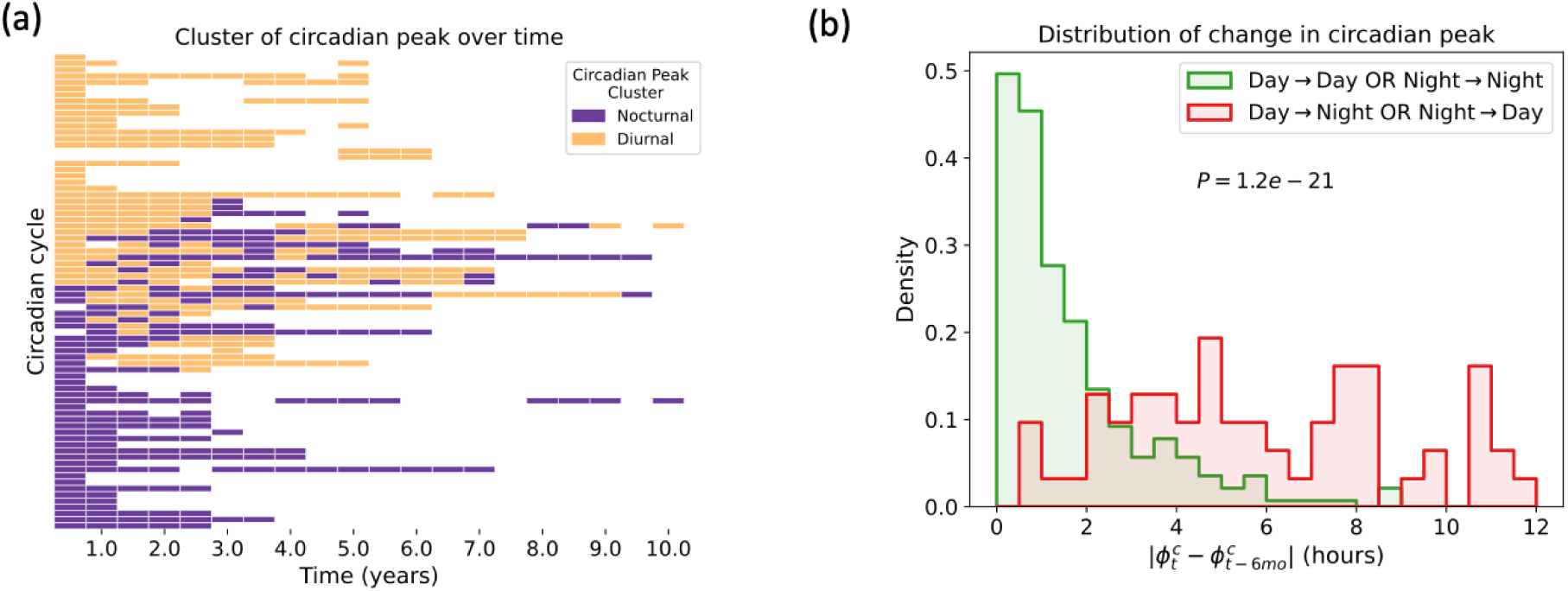
Noctural and diurnal clusters of circadian cycle peaks. (a) Cluster membership of circadian cycle peak hour over the entire data for different participants and hemispheres. (b) Magnitude of angular change in peak between successive 6-month windows is larger for inter-cluster transitions compared to within-cluster transitions. Abbreviations: 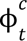 - peak phase of circadian cycle *c* in epoch *t*.

### Evaluating the prediction of ongoing seizure reduction using stimulation features

**Figure S7:**
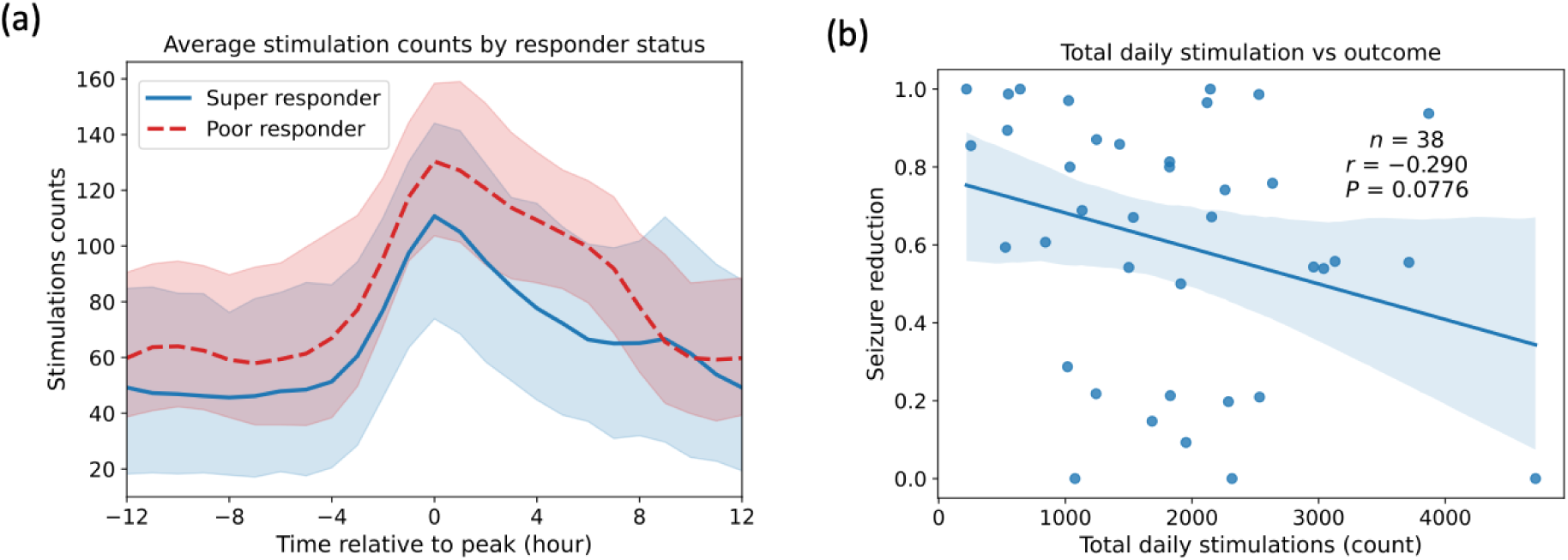
Stimulation counts are not associated with seizure reduction. (a) Circadian profile of hourly stimulation counts for super responders (seizure reduction > 0.9) and poor responders (seizure reduction < 0.5). Stimulation profiles were aligned such that maximum stimulation counts occurred at 0h within each participant. Counts were not significantly different between the groups at any hour. (b) Total stimulation, measured as the sum across hours of the circadian stimulation counts per participant, was not associated with average seizure reduction.

We examined whether stimulation features with circadian features could predict longitudinal, rather than average, seizure reduction. Normalized hourly stimulation was computed within non-overlapping six-month epochs from implantation. Four stimulation-derived features were extracted per epoch: SPA(7AM–11AM), SPA(11AM–7AM), 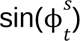, and 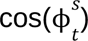. Epochs with circadian and stimulation features were time-matched to the nearest subsequent clinic visit within a year. If multiple epochs were matched to the same clinic visit, then only the most recent one was considered. Multivariate regression models incorporating these features were trained with RFE and evaluated using LOPO cross-validation (Fig. S8).

**Figure S8:**
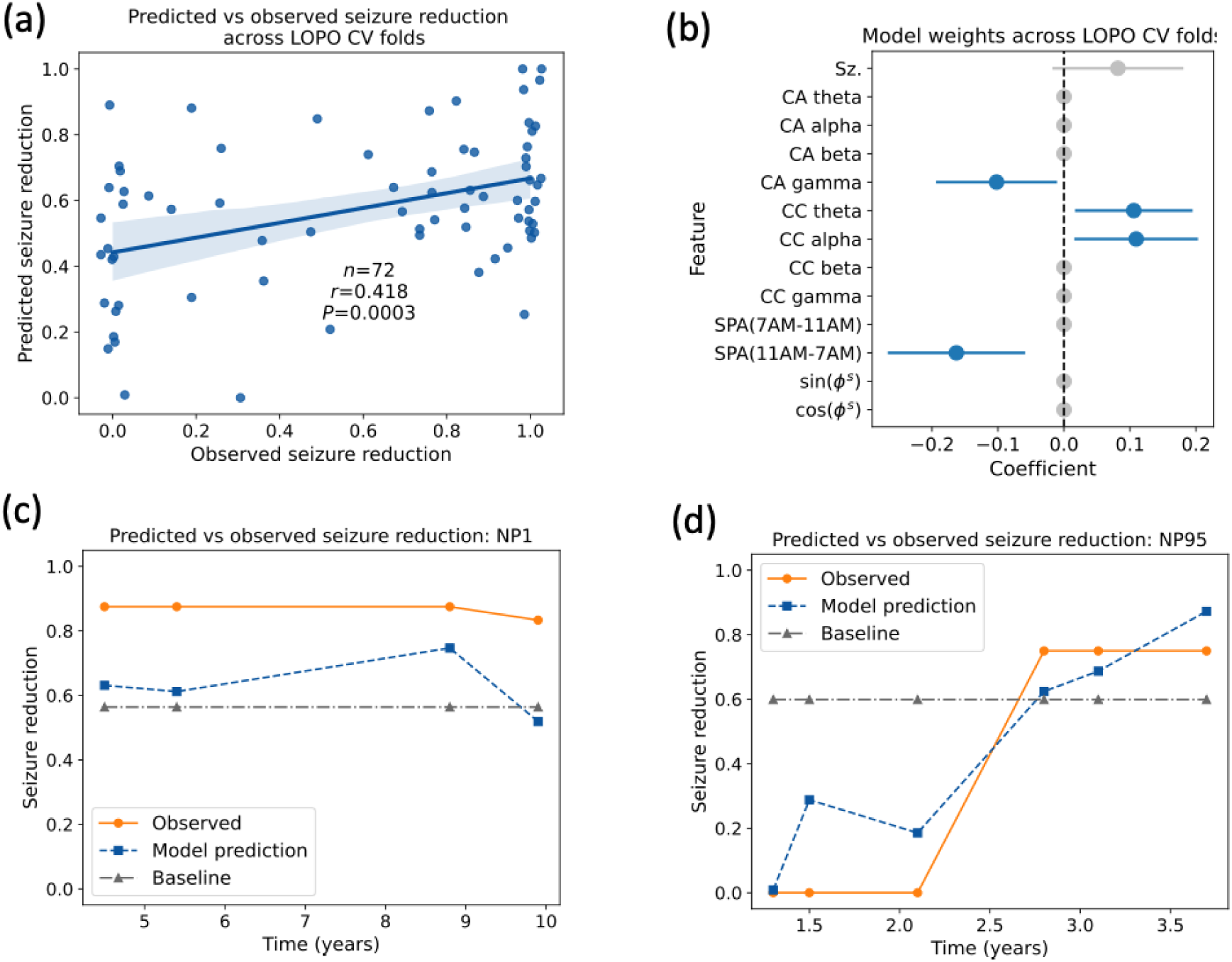
Outcome prediction from time-matched circadian and stimulation features. (a) LOPO cross-validated model predictions for seizure reduction based on time-matched circadian and stimulation features, along with log of baseline seizure rate. Random jitter of 0.03 was added to x-axis for visualization only. (b) Weights of prediction models across cross-validation folds. Features eliminated during RFE are set to 0 (shown in shaded gray). (c, d) Observed and predicted seizure reductions for two example participants. Participants were in the test-set in their respective folds. Baseline corresponds to population mean seizure reduction in the training data.

### Kuramoto model performance evaluation

We compared the full Kuramoto model with baseline models in their ability to predict the circadian cycle peak phases at the end of six-month epochs for all participants. Prediction error was defined as the total absolute difference between observed and model-predicted peak hours across hemispheres. The full model achieved significantly lower prediction error than either baseline (Wilcoxon signed-rank test (paired test); *n1 = n2* = 165; *P* < 4.9e−5) (Fig. S9b). The median error of the full model was 0.02 hours; however, the error distribution was bimodal, with a secondary mode near 10.97 hours, indicating that a subset of samples was not well captured by the model or the coupling constant estimation procedure.

To assess the generalizability of the Kuramoto model across clinical outcomes, we stratified epochs by time-matched seizure reduction (seizure reduction < 0.5 versus > 0.9) and compared prediction errors between groups, separately for each model variant. The full Kuramoto model showed no significant difference in prediction error between outcome strata (Fig. S9c), indicating that its explanatory capacity was not contingent on a particular therapeutic response.

**Figure S9:**
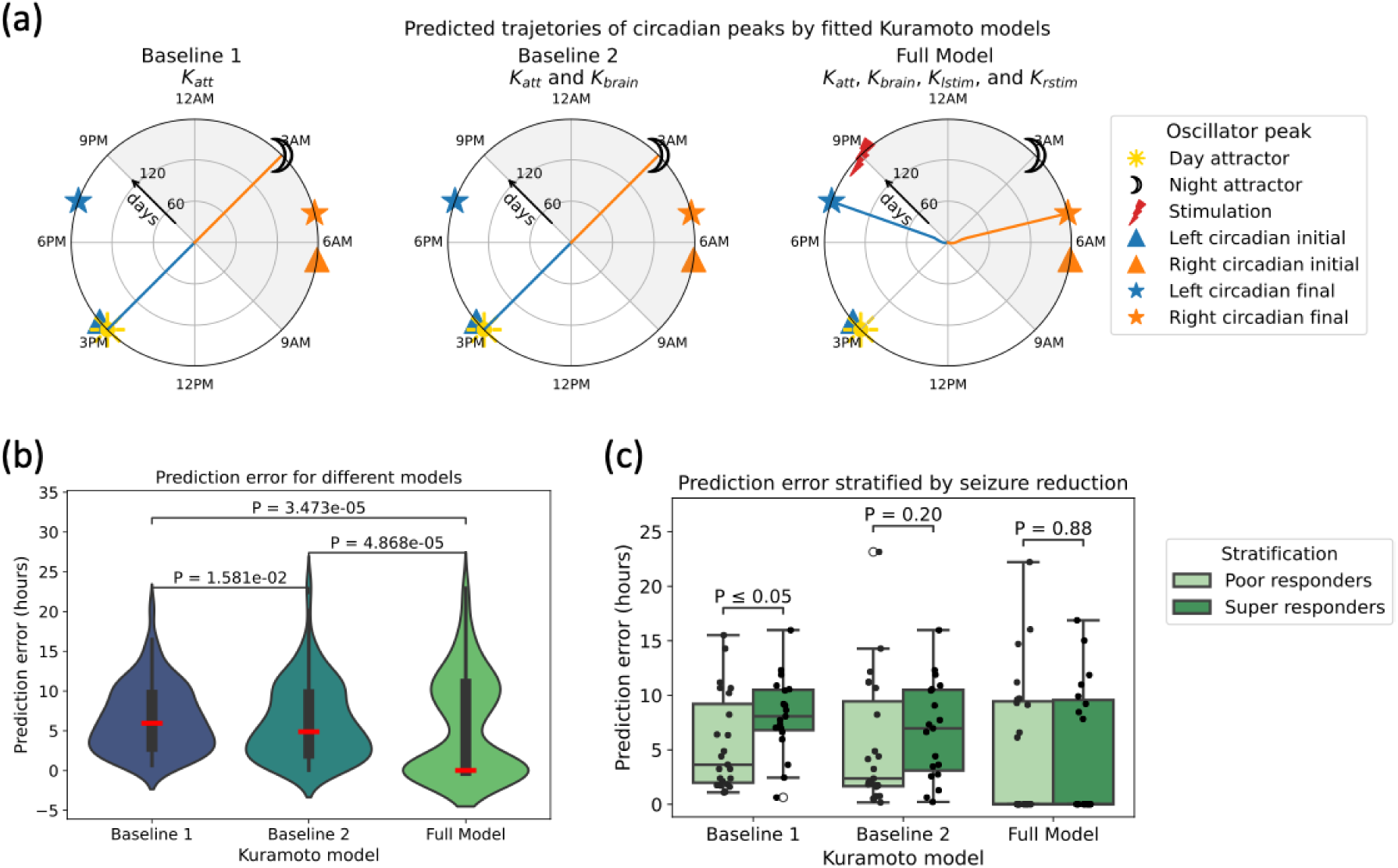
Kuramoto model performance evaluation. (a) Predicted trajectory of circadian peaks over six months for a real sample for Kuramoto models with different coupling constants. Same example as in Figure 6c, but with unscaled, empirically estimated coupling values. (b) Distribution of prediction error of Kuramoto model across six-month epochs for models with different coupling constants. Median is shown in red. (c) Distribution of prediction error across six-month epochs stratified by observed responder status for Kuramoto models with different coupling constants.

### Prediction of Kuramoto model coupling constants for the in-silico framework

Separate predictive models were developed for each coupling constant. A multivariate regression model with RFE was trained to predict *K_brain_* from features of the initial circadian cycles and stimulation. This model achieved significantly better-than-chance performance (*n* = 79, *r* = 0.505, *P* = 2e-6) (Fig. S10a,b). To predict stimulation–cycle coupling strength (*K_lstim_* and *K_rstim_*) as a function of stimulation peak hour, hierarchical regression models were trained using the initial circadian cycle peak phase and amplitude alongside stimulation features. LOPO cross-validation demonstrated that these models predicted empirically estimated coupling strengths significantly better than chance (*K_lstim_* and *K_rstim_* : *n* = 164, *r* = 0.306, *P* = 6.8e−5; Fig. 7b). Stimulation profile area and initial cycle phase were the most influential predictors, consistent with earlier analyses linking SPA to seizure reduction (Fig. S10c). Notably, the ability to predict Kuramoto coupling constants from biologically interpretable features provides additional support for the mechanistic validity of this framework. If the coupling parameters captured arbitrary variance rather than genuine physiological interactions, they would not be predictable from features with established biological relevance.

**Figure S10:**
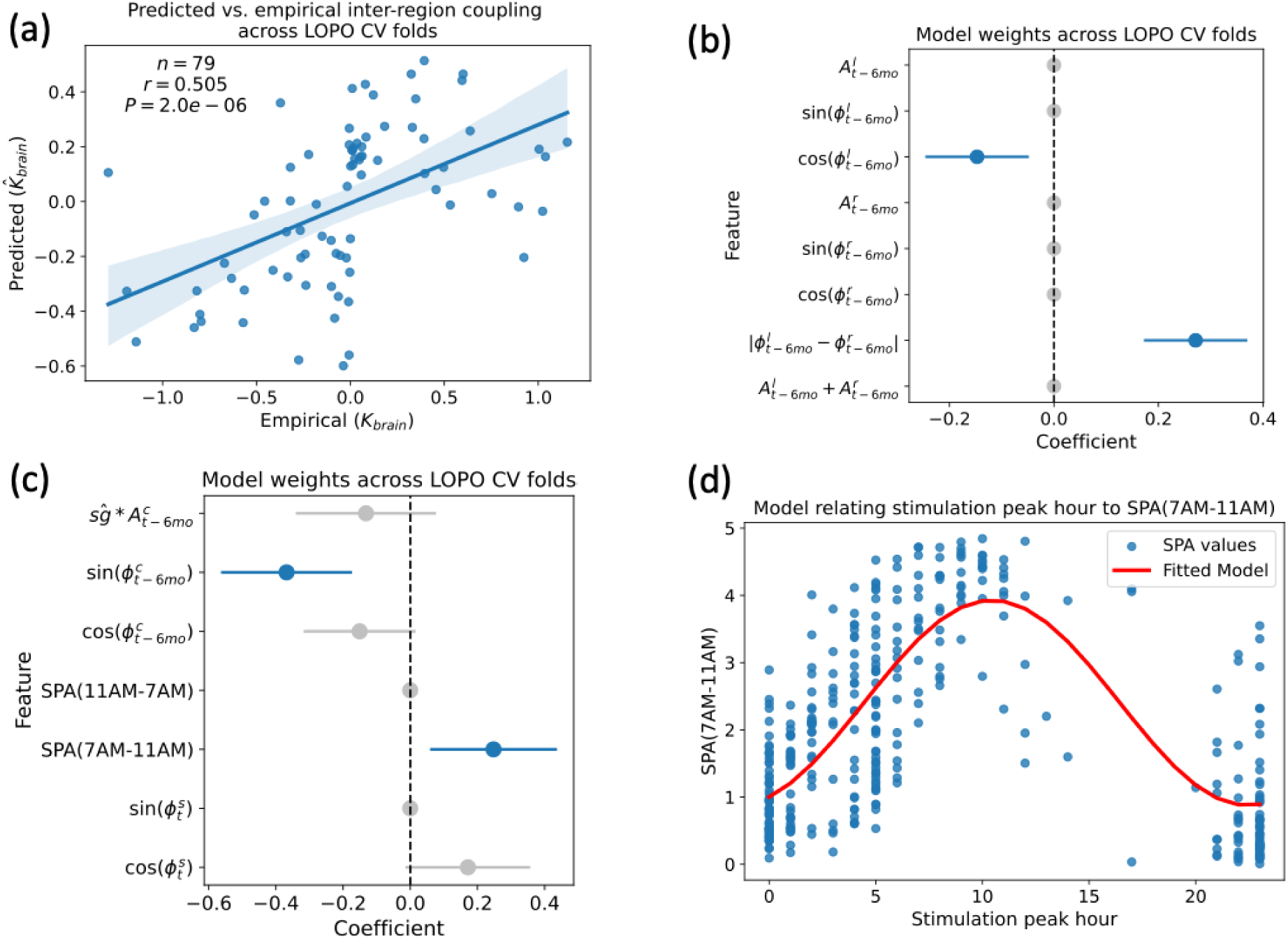
Prediction of coupling constants using circadian and stimulation features. (a) Cross-validated model predictions for inter-region coupling (*K_brain_*) based on circadian cycle features at the initial time point (*t* − 6*mo*). (b) Weights of *K_brain_* prediction models across cross-validation folds. Features eliminated during RFE are set to 0 (shown in shaded gray). (c) Weights of stimulation–cycle coupling (*K_lstim_* and *K_rstim_*) prediction models across cross-validation folds. Features eliminated during RFE are set to 0. (d) Sinusoidal model captures the relationship between peak hour of stimulation and SPA(7AM-11AM). This model was used to estimate SPA(7AM-11AM) for virtual stimulation with a given peak hour for the downstream prediction of stimulation–cycle coupling. Notation: 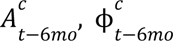–circadian cycle amplitude and peak phase at the start of the six-month epoch, where relevant, *c* is replaced by *l* (left) or *r* (right); 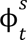–stimulation peak phase during the six-month epoch; 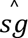- sing.

### Validation of in silico framework

For a given peak hour of virtual stimulation, the Kuramoto model-based *in silico* framework was used to predict the circadian cycle peaks trajectory over six months. The terminal phase of the circadian cycles at six months was used to obtain the predicted circadian concordance induced to the given virtual stimulation. By exploring all 24 candidate virtual stimulation peak hours across all samples (epochs), we computed the average circadian concordance as a function of stimulation peak hour across the population (Fig. S11b). Concordance was highest when stimulation peaked between 5 AM and 8 AM. The local minima observed near 3 AM and 3 PM reflect the influence of the attractor oscillators at those phases. When stimulation aligns with an attractor, cycles are pulled toward that attractor independently, driving concordance toward either 0 or 1 depending on whether the two cycles are attracted to the same or opposite attractors. These results indicate that stimulation peak hour meaningfully influences population-level circadian concordance, with an early-morning delivery window being most conducive to bilateral network alignment.

To further validate the *in silico* framework against real data, we examined the relationship between stimulation profile area and concordance across the 24 simulated stimulation peak hours. SPA(11AM–7AM) was negatively correlated with model-predicted circadian concordance (Mixed effects model; *n* = 3853, β = −0.015, *P* = 0.041) (Fig. S11c). This relationship emerged from the Kuramoto model dynamics without any direct exposure to the empirical association between SPA and seizure reduction, thus, supporting the validity of the *in silico* framework.

**Figure S11:**
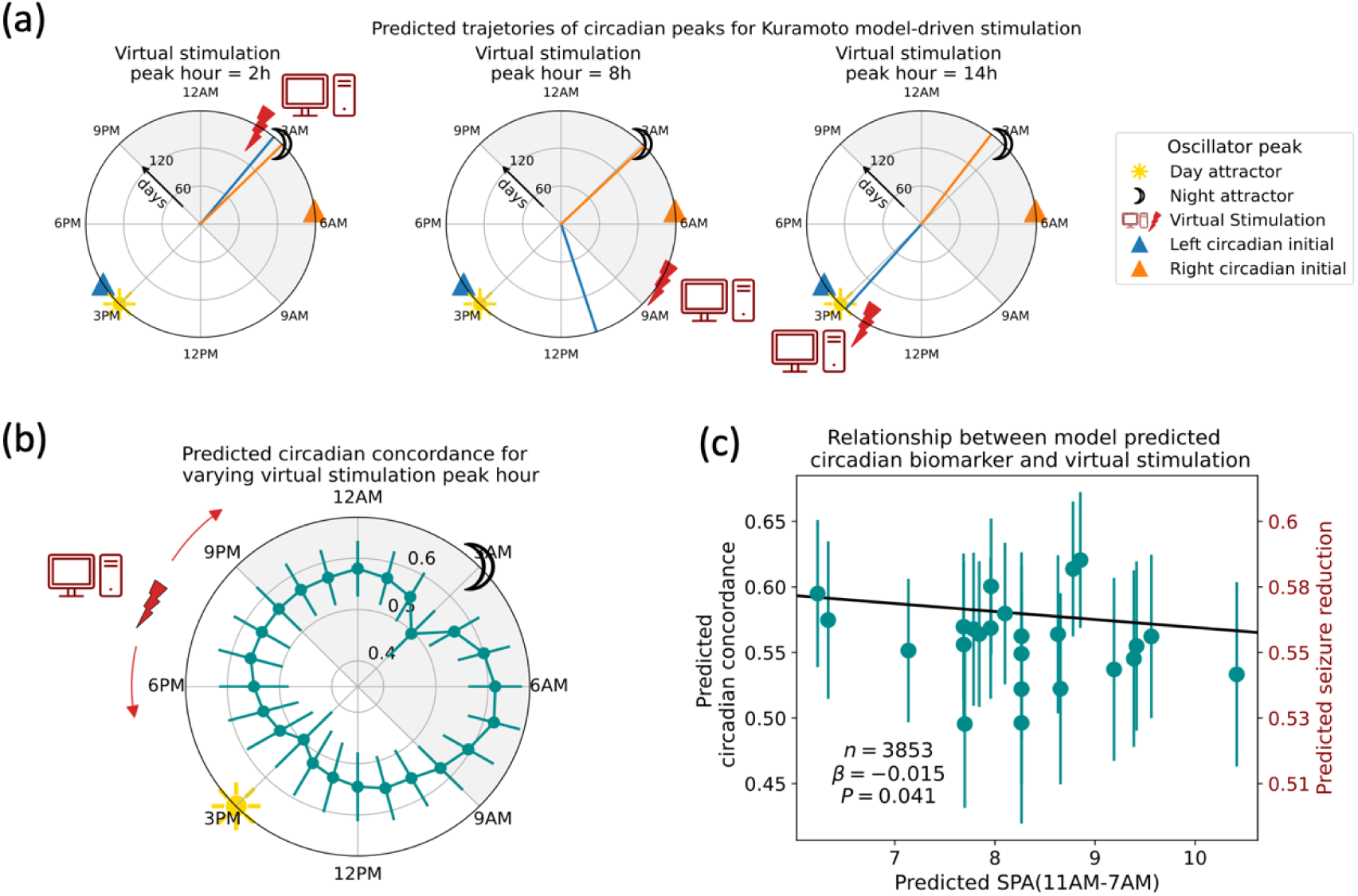
Validation of *in silico* framework for optimizing stimulation. (a) Effect of varying the peak hour of virtual stimulation on circadian cycles. Subplots show, for a real sample, the changes in Kuramoto model-predicted trajectory of circadian peaks over six months for different virtual stimulation peak hours (shown with a computer icon and red lightning bolt). Same example as in Figure 7c, but with unscaled coupling values obtained from coupling prediction pipeline. (b) Average predicted circadian concordance induced by virtual stimulation as the stimulation’s peak hour was varied over 24 hours. Average was taken across all epochs. (c) Circadian concordance from virtual stimulation was associated with SPA(11AM-7AM). Mixed effect model was used to account for repeated measures within participants.

## References

1. Baud, M. O. et al. Multi-day rhythms modulate seizure risk in epilepsy. Nat. Commun. 9, 88–88 (2018).

2. Zhang, X. et al. Rhythms in longitudinal thalamic recordings are linked to seizure risk. Epilepsia 67, 2559–2571 (2026).

3. Chiang, S. et al. Evidence of state-dependence in the effectiveness of responsive neurostimulation for seizure modulation. Brain Stimulat. 14, 366–375 (2021).

4. Chiang, S. et al. State-dependent effects of responsive neurostimulation depend on seizure localization. Brain 148, 521–532 (2025).

5. Dixon, T. C. et al. Movement-responsive deep brain stimulation for Parkinson’s disease using a remotely optimized neural decoder. *Nat*. Biomed. Eng. 10, 110–124 (2025).

6. Karoly, P. J. et al. Cycles in epilepsy. Nat. Rev. Neurol. 17, 267–284 (2021).

7. Charlebois, C. M. et al. Circadian changes in aperiodic activity are correlated with seizure reduction in patients with mesial temporal lobe epilepsy treated with responsive neurostimulation. Epilepsia 65, 1360–1373 (2024).

8. Cho, C.-H. Molecular mechanism of circadian rhythmicity of seizures in temporal lobe epilepsy. Front. Cell. Neurosci. 6, (2012).

9. Zamora, M. et al. DyNeuMo Mk-1: Design and pilot validation of an investigational motion-adaptive neurostimulator with integrated chronotherapy. Exp. Neurol. 351, 113977 (2022).

10. Nair, D. R. et al. Nine-year prospective efficacy and safety of brain-responsive neurostimulation for focal epilepsy. Neurology 95, e1244–e1256 (2020).

11. Rao, V. R. & Rolston, J. D. Unearthing the mechanisms of responsive neurostimulation for epilepsy. Commun. Med. 3, 1–6 (2023).

12. Khambhati, A. N., Shafi, A., Rao, V. R. & Chang, E. F. Long-term brain network reorganization predicts responsive neurostimulation outcomes for focal epilepsy. Sci. Transl. Med. 13, eabf6588 (2021).

13. Kokkinos, V., Sisterson, N. D., Wozny, T. A. & Richardson, R. M. Association of Closed-Loop Brain Stimulation Neurophysiological Features With Seizure Control Among Patients With Focal Epilepsy. JAMA Neurol. 76, 800 (2019).

14. Khambhati, A. N., Chang, E. F., Baud, M. O. & Rao, V. R. Hippocampal network activity forecasts epileptic seizures. Nat. Med. 30, 2787–2790 (2024).

15. Anderson, D. N. et al. Closed-loop stimulation in periods with less epileptiform activity drives improved epilepsy outcomes. Brain 147, 521–531 (2024).

16. Provenza, N. R. et al. Disruption of neural periodicity predicts clinical response after deep brain stimulation for obsessive-compulsive disorder. Nat. Med. 30, 3004–3014 (2024).

17. Dary, Z. et al. Circadian modulation of aperiodic background and oscillatory activity as a biomarker of seizure burden in Lennox–Gastaut syndrome. Epilepsia epi.70457 (2026) doi:10.1002/epi.70457.

18. Eliashiv, D. et al. Postapproval Study for Brain-Responsive Neurostimulation for Drug-Resistant Focal Epilepsy: Three-Year Efficacy and Interim Safety Results. Neurology 106, e214875 (2026).

19. Razavi, B. et al. Real-world experience with direct brain-responsive neurostimulation for focal onset seizures. Epilepsia 61, 1749–1757 (2020).

20. Baud, M. O., Bernard, C., Frauscher, B., Karoly, P. J. & Rao, V. R. Timing is everything: Expert opinion on researching epilepsy rhythms by the ILAE Task Force on Chronobiology. Epilepsia 67, 1544–1554 (2026).

21. Bradley, C., Nydam, A. S., Dux, P. E. & Mattingley, J. B. State-dependent effects of neural stimulation on brain function and cognition. Nat. Rev. Neurosci. 23, 459–475 (2022).

22. Ruoff, L. et al. Sleep disruption is not observed with brain-responsive neurostimulation for epilepsy. Epilepsia Open 5, 155–165 (2020).

23. Peter-Derex, L., et al. Sleep Disruption in Epilepsy: Ictal and Interictal Epileptic Activity Matter. Ann. Neurol. 88, 907–920 (2020).

24. Bower, M. R. et al. Reactivation of seizure-related changes to interictal spike shape and synchrony during postseizure sleep in patients. Epilepsia 58, 94–104 (2017).

25. Klinzing, J. G., Niethard, N. & Born, J. Mechanisms of systems memory consolidation during sleep. Nat. Neurosci. 22, 1598–1610 (2019).

26. Chapeton, J. I., Haque, R., Wittig, J. H., Inati, S. K. & Zaghloul, K. A. Large-Scale Communication in the Human Brain Is Rhythmically Modulated through Alpha Coherence. Curr. Biol. 29, 2801–2811.e5 (2019).

27. Warren, C. P. et al. Synchrony in Normal and Focal Epileptic Brain: The Seizure Onset Zone is Functionally Disconnected. J. Neurophysiol. 104, 3530–3539 (2010).

28. Thornton, C. et al. Diminished circadian and ultradian rhythms of human brain activity in pathological tissue in vivo. Nat. Commun. 15, 8527 (2024).

29. Khambhati, A. N. et al. Dynamic Network Drivers of Seizure Generation, Propagation and Termination in Human Neocortical Epilepsy. PLOS Comput. Biol. 11, e1004608 (2015).

30. Rashid, S., Pho, G., Czigler, M., Werz, M. A. & Durand, D. M. Low frequency stimulation of ventral hippocampal commissures reduces seizures in a rat model of chronic temporal lobe epilepsy. Epilepsia 53, 147–156 (2012).

31. Toprani, S. & Durand, D. M. Fiber tract stimulation can reduce epileptiform activity in an in-vitro bilateral hippocampal slice preparation. Exp. Neurol. 240, 28–43 (2013).

32. Hadar, P. N., Zelmann, R., Salami, P., Cash, S. S. & Paulk, A. C. The Neurostimulationist will see you now: prescribing direct electrical stimulation therapies for the human brain in epilepsy and beyond. Front. Hum. Neurosci. 18, (2024).

33. Makhoul, G. S. et al. Pathway to Adaptive Neuromodulation: Modeling Epileptogenic Brain States Using SEEG. Epilepsy Curr. 15357597261422447 (2026) doi:10.1177/15357597261422447.

34. McCarthy, P. M. & Burdette, D. E. Responsive neurostimulation (RNS) programming. Neurotherapeutics 23, e00944 (2026).

35. Herzel, H., Lilienthal, S., Hammerstein, P. & Del Olmo, M. Theoretical chronobiology of circadian timing—gold mine or minefield? Npj Biol. Timing Sleep 3, 32 (2026).

36. Depannemaecker, D., Ezzati, A., Wang, H. E., Jirsa, V. & Bernard, C. From phenomenological to biophysical models of seizures. Neurobiol. Dis. 182, 106131 (2023).

37. Oganesian, L. L. & Shanechi, M. M. Brain–computer interfaces for neuropsychiatric disorders. Nat. Rev. Bioeng. 2, 653–670 (2024).

38. El Houssaini, K., Bernard, C. & Jirsa, V. K. The Epileptor Model: A Systematic Mathematical Analysis Linked to the Dynamics of Seizures, Refractory Status Epilepticus, and Depolarization Block. eneuro 7, ENEURO.0485-18.2019 (2020).

39. Jirsa, V. et al. Personalised virtual brain models in epilepsy. Lancet Neurol. 22, 443–454 (2023).

40. Wang, H. E. et al. Virtual brain twins for stimulation in epilepsy. Nat. Comput. Sci. 5, 754–768 (2025).

41. Buchin, A., Chizhov, A., Huberfeld, G., Miles, R. & Gutkin, B. S. Reduced Efficacy of the KCC2 Cotransporter Promotes Epileptic Oscillations in a Subiculum Network Model. J. Neurosci. 36, 11619–11633 (2016).

42. Yang, Y. et al. Modelling and prediction of the dynamic responses of large-scale brain networks during direct electrical stimulation. *Nat*. Biomed. Eng. 5, 324–345 (2021).

43. Saboo, K. V. et al. State space modeling of multidien cyclical progression of epilepsy.

44. Aboumatar, S. et al. Electrographic seizures on responsive neurostimulation: An early and objective measure of response to cenobamate. Epilepsy Res. 218, 107647 (2025).

45. Matsuda, T. & Komaki, F. Multivariate Time Series Decomposition into Oscillation Components. Neural Comput. 29, 2055–2075 (2017).

46. Von Wegner, F., Bauer, S., Rosenow, F., Triesch, J. & Laufs, H. EEG microstate periodicity explained by rotating phase patterns of resting-state alpha oscillations. NeuroImage 224, 117372 (2021).

47. 47. Feys, O. et al. Virtual Responsive Neurostimulation Implantation: From Intracranial Connectivity to Optimized Lead Placement. Preprint at 10.64898/2026.06.17.26355892 (2026).

48. Liou, J. et al. Role of inhibitory control in modulating focal seizure spread. Brain 141, 2083–2097 (2018).

49 . The Astropy Collaboration et al. The Astropy Project: Sustaining and Growing a Community-oriented Open-source Project and the Latest Major Release (v5.0) of the Core Package*. Astrophys. J. 935, 167 (2022).

50. VanderPlas, J. T. Understanding the Lomb–Scargle Periodogram. Astrophys. J. Suppl. Ser. 236, 16 (2018).

51. Balzekas, I. et al. Method for cycle detection in sparse, irregularly sampled, long-term neuro-behavioral timeseries: Basis pursuit denoising with polynomial detrending of long-term, inter-ictal epileptiform activity. PLOS Comput. Biol. 20, e1011152 (2024).

52. Seabold, S. & Perktold, J. Statsmodels: Econometric and Statistical Modeling with Python. in 92–96 (Austin, Texas, 2010). doi:10.25080/Majora-92bf1922-011.

53. Bauer, L. G. et al. Quantification of Kuramoto Coupling Between Intrinsic Brain Networks Applied to fMRI Data in Major Depressive Disorder. Front. Comput. Neurosci. 16, 729556 (2022).

54. Zheng, S. et al. Kuramoto Model-Based Analysis Reveals Oxytocin Effects on Brain Network Dynamics. Int. J. Neural Syst. 32, 2250002 (2022).

